# The Effects of Gender Affirming Hormone Treatment on Transgender Women’s Musculoskeletal Health: A Systematic Review and Meta-Analysis

**DOI:** 10.1101/2025.05.21.25327924

**Authors:** Andrew Brown, Stephanie Montagner-Moraes, Jatinder Singh, Laura Charlton, James Barrett, Blair R. Hamilton

## Abstract

**Background:** Gender-affirming hormone treatment (GAHT) aligns transgender women’s hormone profiles with their gender identity, alleviating gender dysphoria by inducing feminising changes. However, the effects of GAHT on musculoskeletal health, particularly bone mineral density (BMD), require ongoing evaluation. A previous meta-analysis showed GAHT had a small effect on lumbar spine (LS) BMD, but more recent studies and updated methodologies justify a new synthesis. **Methods:** A systematic review and meta-analysis were conducted using studies published in English up to 31/07/2024, identified via three electronic databases, cross-referencing, and expert review. Primary outcomes were changes in femoral neck (FN), LS, and total hip (TH) BMD. Secondary outcomes included changes in body composition. Standardised effect sizes (Hedges’ g) were pooled using the inverse heterogeneity (IVhet) model. **Results:** GAHT was associated with a significant increase in FN BMD (g = 0.13 [0.05, 0.20], *p* = 0.00). Significant gains were also observed in fat mass (FM) (g = 0.52), BMI (g = 0.16), and body fat percentage (BF%) (g = 0.79), while reductions were found in fat-free mass (FFM) (g = -0.21) and thigh muscle cross-sectional area (mCSA) (g = -1.02). **Conclusion:** GAHT maintains FN BMD in transgender women with increased FM and reduced fat-free mass. The heterogeneous nature of several outcomes and the absence of empirical data on ageing transgender women require further research and clinical monitoring of bone health (such as High-resolution peripheral quantitative computed tomography) and muscle health to clarify the long-term implications of GAHT, particularly as transgender women age.

## Introduction

Transgender women often experience psychological distress due to a mismatch between their assigned gender at birth and their experienced gender, a condition medically diagnosed as gender incongruence of adulthood [1, 2]. Approximately 3% of the global population, equating to around 241 million people [3] identify as transgender. Many of these individuals undergo medical interventions, including gender-affirming hormone treatment (GAHT) and gender-affirming surgery, to align their physical characteristics with their gender identity [4, 5]. The demand for transgender and gender diverse health services has notably increased in recent years across several European nations and first-world nations globally [6–8] reflecting a growing recognition and acceptance of gender diversity.

For transgender women (individuals assigned male at birth who identify as female), GAHT usually involves a combination of oestrogen and anti-androgens, which have been shown to reduce distress and increase the quality of life in transgender women [9–11]. Oestrogen promotes feminising effects such as breast development, softening of skin, reduction in muscle mass, and redistribution of body fat to hips and thighs [12]. Anti-androgens work by reducing the effects of testosterone, further facilitating the feminisation process [4, 12, 13].

Androgens like testosterone are essential for maintaining skeletal homeostasis [14–16] And oestrogen has a well-established positive effect on bone homeostasis [17–20]. These physiological contexts suggest that alterations of sex steroids in transgender women can significantly alter their musculoskeletal health. Consequently, the implications of sex steroids on bone health must be carefully considered in transgender women, especially in the context of GAHT. However, the effects of GAHT on their muscle health are less understood.

A previous meta-analysis by Singh-Ospina [21] investigated the effects of GAHT on the bone health of transgender individuals. In transgender women, the authors found an increase in lumbar spine (LS) bone mineral density (BMD) at both 12 and 24 months (Effect Size (ES) = 12 months 0.04 [-0.03, 0.06]), 24 months 0.06 [0.04, 0.08]), while finding no change in femoral neck (FN) BMD (ES = 12 months 0.02 [0.00, 0.03]), 24 months 0.06 [0.00, 0.03]). The authors concluded that in transgender women, GAHT was associated with an increase in BMD at the LS While the results reported by Singh-Ospina [21] are noteworthy, they were limited to only thirteen trials, observational studies, and case series published up to April 2015, lacking an assessment of BMD using quantitative computed tomography (QCT) [21], and do not provide an assessment of muscle health. Since Singh-Ospina [21], additional studies have been published [22–24] and more robust methods for the undertaking and interpretation of meta-analytic results have been developed [25–28]. Furthermore, to the best of the authors’ knowledge, no previous systematic reviews with meta-analysis or original systematic reviews with meta-analysis have been conducted on the effects of GAHT on overall musculoskeletal health in transgender women since the original analysis. Finally, using previously developed guidelines for when to update a systematic review, it was decided that an updated review on this topic was needed [29]. Thus, given 1) the updated preferred reporting items for systematic reviews and meta-analyses (PRISMA) statement [30]2) the potential effects of GAHT on transgender women’s musculoskeletal health outside of BMD, 3) the lack of recent meta-analytic work in this area, 4) the use of more robust methods for conducting meta- analytic research [25–28] and 5) decision tree analysis of when to update a systematic review [29] we aim to update and expand on the systematic review with a meta-analysis by Singh-Ospina [21], whereby we will examine the effects of GAHT on transgender women’s musculoskeletal health.

## Materials and Methods Study Eligibility Criteria

As this meta-analysis aims to update the Singh-Ospina et al. Singh-Ospina [21] meta- analysis, the same *a priori* inclusion criteria were employed (**Table 1**), with additional studies identified from 07/04/2015. Studies that do not meet the criteria outlined in Table 1 were excluded from the analysis. This meta-analysis adhered to the PRISMA guidelines [31]. The protocol was preregistered in PROSPERO (trial registration number: CRD42024573102 [32]).

**Table 1.**
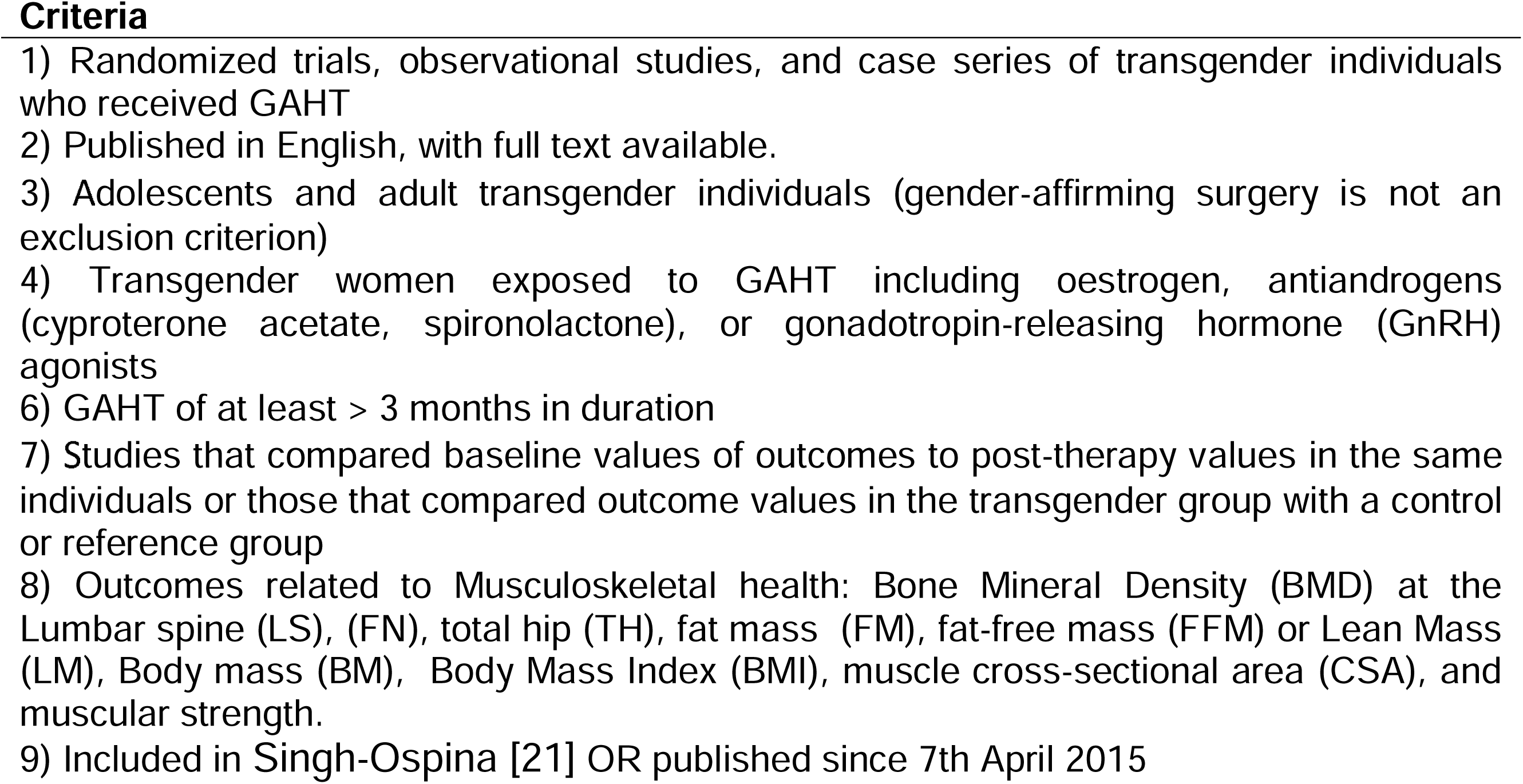
Study Eligibility Criteria Criteria

## Data Sources

Studies published up to 31^st^ July 2024 were retrieved from three electronic sources (PubMed, Embase, SportDiscus). Keywords relevant to all searches included “transgender,” “bone,” and “muscle.” A final database search was conducted on 1^st^ December 2024. Based on PRISMA guidelines [31], an example of the search strategy is supplied in the supplementary material [33]. The last author (BRH) conducted all electronic database searches. In addition to electronic database searches, cross-referencing from retrieved studies was also conducted.

## Study Records and Selection

All studies were imported into EndNote (EndNote 20.6, Clarivate Analytics, USA), and the last author (BRH) removed duplicates electronically and manually. A copy of the reference database was then provided to the first author (AB) and the second (SMM) for dual screening. Both authors (AB and SMM) screened studies independently. The screeners were not blinded to journal titles or the study authors/affiliations. Reasons for exclusion were coded based on one or more of the following: 1) inappropriate population, 2) inappropriate intervention, 3) inappropriate comparison(s), 4) inappropriate outcome(s), 5) inappropriate study design or 6) other. On completion, the screeners met to discuss their selections and reconcile any discrepancies by consensus. If an agreement cannot be achieved, the last author (BRH) provided a recommendation. The agreement rate, before the reconciliation of any discrepancies, was calculated using Cohen’s κ statistic [34]. The precision of searches was calculated as the number of studies included divided by the number of studies screened (less duplicates) [35]. The number needed to screen (NNS) was calculated by taking the reciprocal of the precision [35].

## Data Abstraction

Before data abstraction, an electronic codebook was developed by the last author (BRH) and provided to the joint first authors (AB/SMM). The extracted data were coded based on the following major categories: 1) study characteristics (e.g., author, journal, year, etc.), 2) participant characteristics (e.g., age, height, mass, etc.), 3) intervention details (e.g., type, length, frequency, etc.), and 4) outcome characteristics (e.g., sample sizes, baseline/post-GAHT means and SD, etc.). A copy of the electronic codebook is available in the supplementary material [33].

The first (AB and SMM) and last (BRH) authors extracted all data independently of one another before meeting to resolve any discrepancies by consensus. If an agreement was not achieved, the last author (BRH) provided a recommendation. Before this, the overall agreement rate was assessed by Cohen’s κ statistic [34].

### Outcome Measures

*A priori* primary outcome measures were changes in bone health parameters such as Total Hip (TH), Femoral Neck (FN) and Lumbar Spine (LS) bone mineral density (BMD) measured by Dual-Energy X-Ray Absorptiometry (DXA), Dual-Photon Absorptiometry (DPA), or Quantitative Computed Tomography (QCT). Secondary, *a priori* outcomes included changes in body mass (BM), body mass index (BMI), Lean body mass (LM) or fat-free mass (FFM), fat mass (FM), muscle cross-sectional area (CSA) and muscular strength. Obtaining missing data was attempted for all primary and secondary outcome measures if assessed by a study and the data provided proves inadequate to calculate an effect size. The last author (BRH) contacted the study’s corresponding author three times via email, with one week between each communication. These communications were tracked (e.g., dates, responses, success rates, etc.) to establish the success rate of this process.

### Risk of Bias Assessment

The risk of bias for each study was assessed using the recently revised Cochrane Risk of Bias instrument for non-randomised studies of interventions (ROBINS-I) [36]. Using one or more signalling questions, the ROBINS-I instrument assessed the risk of bias in seven distinct domains: (1) bias arising from confounding, (2) bias in participant selection (3) bias in classification of interventions (4) bias due to deviations from intended interventions, (5) bias due to missing data, (6) bias in the measurement of outcomes and (7) bias in the selection of the reported result. Based on signalling questions, each domain was assessed as either ‘low risk,’ ‘moderate risk,’ ‘serious risk,’ or ’critical risk.’ Based on responses to each domain, the overall risk of bias for each study was assessed as either ‘low risk,’ ‘moderate risk,’ ‘serious risk,’ or ‘‘critical risk.’ We chose to use this risk of bias instrument over the various study quality instruments, including those focused on intervention studies [37, 38] given the difficulty of the latter in differentiating between the quality of reporting and the quality of the conduct of a study [39].

No studies were excluded from the analysis based on the risk of bias assessment [40].Our decision not to exclude on risk of bias was guided by the recognition that the field of transgender musculoskeletal health remains in its early stages, with a relatively limited number of available studies. Excluding all studies with a serious or critical risk of bias would have severely restricted the scope of the analysis and potentially overlooked emerging patterns in the literature. To address this limitation transparently, we conducted sensitivity analyses where possible and explicitly reported the risk of bias assessments alongside study findings. This approach allowed us to highlight the strength of evidence while also acknowledging their limitations. The first (AB) and second (SMM) authors undertook the risk of bias assessment independently of one another, before meeting to resolve any discrepancies by consensus. Where this cannot be achieved, the last author (BRH) provided a recommendation.

### Statistical Analysis Calculation of effect sizes

The *a priori* primary and secondary outcomes for this meta-analysis were calculated using the Hedges standardised mean difference ES, *g*, adjusted for small sample sizes [41]. The *g* for each group was calculated as the mean of the baseline measure or the control/reference group minus the mean of the GAHT intervention group, divided by the pooled and weighted standard deviation. If this information is unavailable, *g* was calculated using procedures described by Follmann *et al*. [42]. For studies reporting multiple post-intervention time points, *g* was calculated based on the baseline and the final time point.

### Effect size pooling

Results were pooled using the inverse heterogeneity (IVhet) model [25], a model which is more robust than the Der Simonian–Laird random effects method employed by Singh- Ospina [21]. Two-tailed z-alpha values <0.05 and non-overlapping 95% confidence intervals will be considered statistically significant. For the longitudinal analysis, outcomes were compared at baseline and at the furthest available time point from baseline within the eligible study. This approach allowed us to assess changes over time within the same individuals. In the cross-sectional analysis, the eligible study must have included a cisgender male comparison group alongside the transgender female group. This comparison was used to evaluate differences between the groups at a single time point, providing a snapshot of the relative outcomes between the two populations.

### Heterogeneity and Inconsistency

For each pooled outcome, heterogeneity was assessed using Q [43], with an alpha level of <0.10 representing statistically significant heterogeneity. Inconsistency was assessed using *I^2^*, an extension of Q. For this meta-analysis, inconsistency was categorised as very low (<25%), low (25-50%), moderate (50-75%) or large (>75%) [43]. Absolute between-study heterogeneity was assessed using tau squared (τ^2^). In addition, influence analysis was conducted by removing each study from our analysis once to examine the effect of that study on the overall findings. Given the expected small sample size, no subgroup or meta-regression analysis was planned a *priori*.

### Meta-biases

Small-study effects (publication bias, etc.) were assessed qualitatively using the Doi plot and quantitatively using the Luis Furuya-Kanamori index (LFK index) [28, 44]. The Doi plot has been suggested to be more intuitive than the funnel plot, and the LFK index is more robust than the commonly used Egger’s regression-intercept test [28, 44]. LFK values within ± 1, greater than ± 1 but within ± 2, and greater than ± 2 were considered to represent no, minor, and major asymmetry [28].

### Strength of evidence

The strength of findings for each outcome was assessed using the most recent version of the Grading of Recommendations Assessment, Development and Evaluation (GRADE) meta-analysis tool [45]. Quality of evidence was assessed across the domains of risk of bias, consistency, directness, precision, and publication bias. Quality was judged as high (further research is very unlikely to change our confidence in the estimate of effect), moderate (further research is likely to have an important impact on our confidence in the estimate of effect and may change the estimate), low (further research is very likely to have an important impact on our confidence in the estimate of effect and is likely to change the estimate), or very low (very uncertain about the estimate of effect) [45]

### Software used for analysis

All data was analysed using MetaXL (version 5.3, Epigear International Pty Ltd). All data is available as supplementary material [46].

## Results

### Study Characteristics

A flow diagram that depicts the search process for study selection is shown in **Figure 1**. A full list of studies is available in the **Supplementary file TransMA_BH_Combined** [46] After identifying 1157 citations, removing 32 studies due to publication before 7th April 2015 and 42 duplicates both electronically and manually, 1083 studies were screened. Of the 1083 studies reviewed, thirty-eight additional studies were included in this meta-analysis, bringing the total number of studies to 55. A post-hoc decision was made to exclude five studies on transgender adolescents under 18 years old [47–51], due to the confounding effects of puberty on musculoskeletal health. The fifty studies represent 47 groups (38 transgender, 9 Cisgender). Of these groups, 2261 participants were transgender women, and 284 participants were comparator cisgender men. The results of transgender men will be published in a separate meta-analysis to provide the reader greater clarity in results and discussion, therefore, only thirty-one studies were included in this meta-analysis.

**Figure 1.**
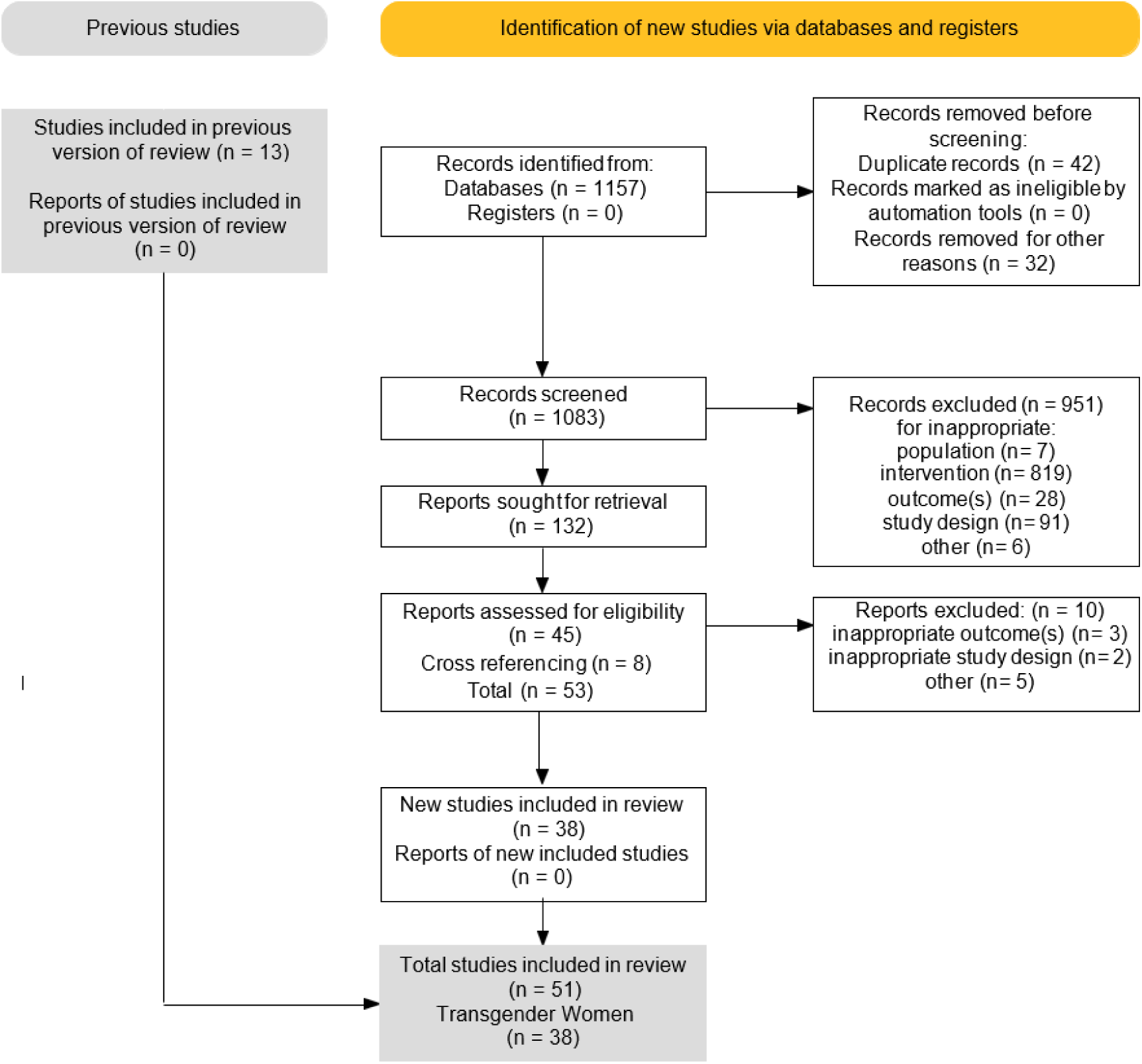
PRISMA Flow Chart

The agreement rate between assessors for inclusion was 0.92 before reconciling any discrepancies, with all discrepancies resolved by consensus between the first and second authors. The major reasons for exclusion were inappropriate population (0.8%), inappropriate intervention (86.2%), inappropriate comparison (0.7%), inappropriate outcome(s) (3.0%), inappropriate study design (9.6%) and other (0.1%). The precision of searches was 12% while the number needed to screen was eight. A list of excluded studies can be found in Supplementary File 1, with a general description of the studies included found in **Table 2**.

**Table 2.**
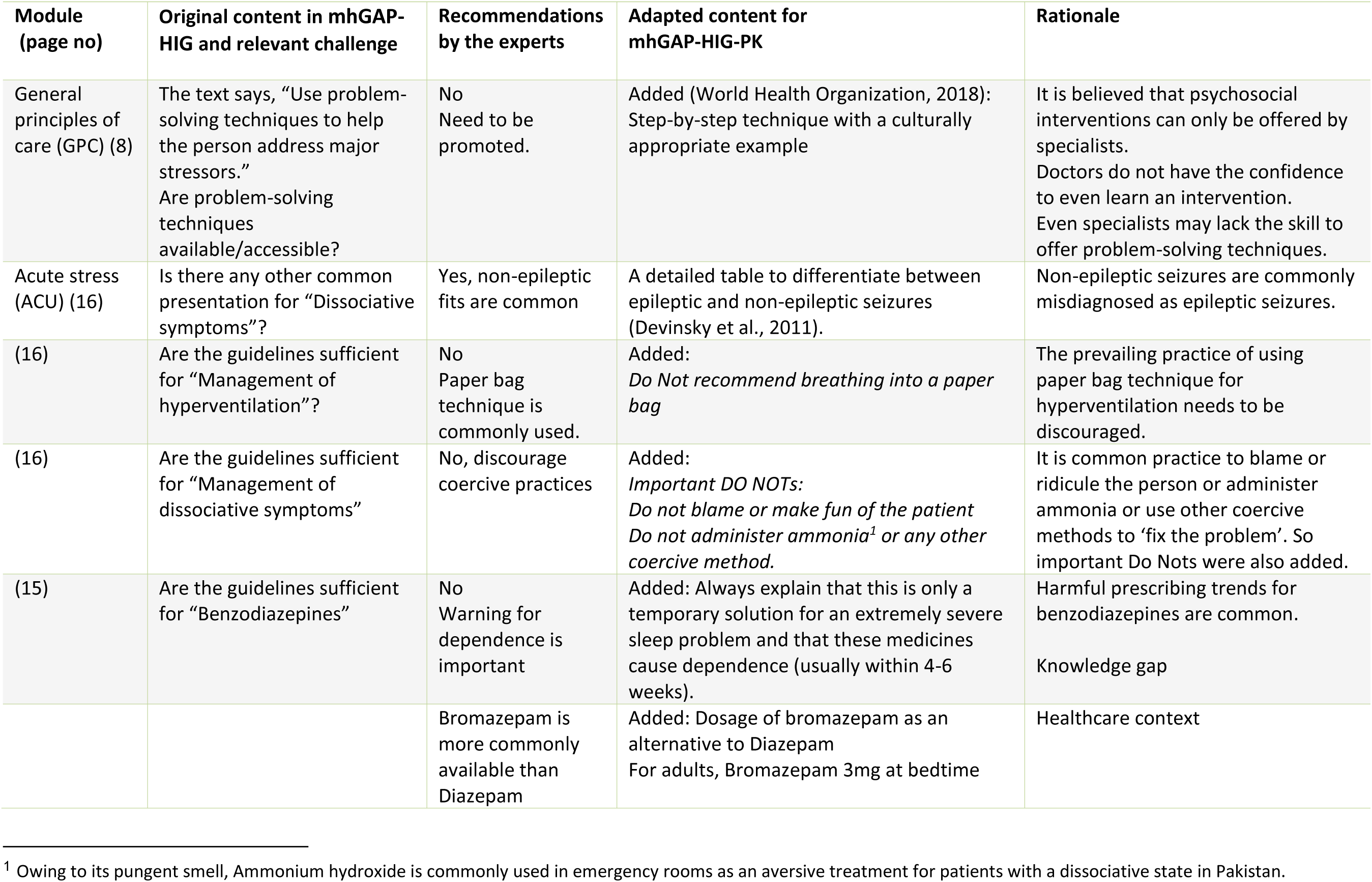
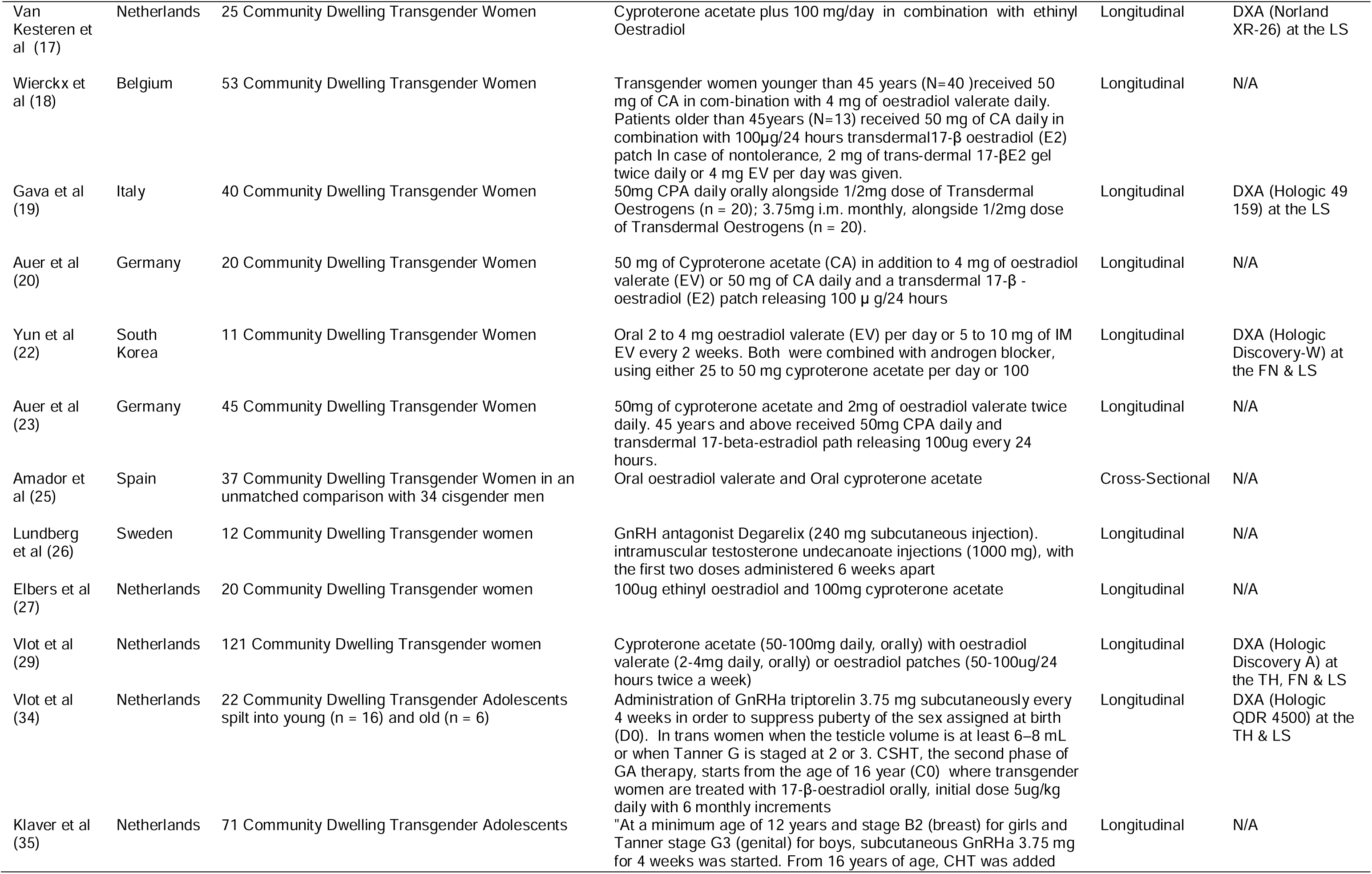
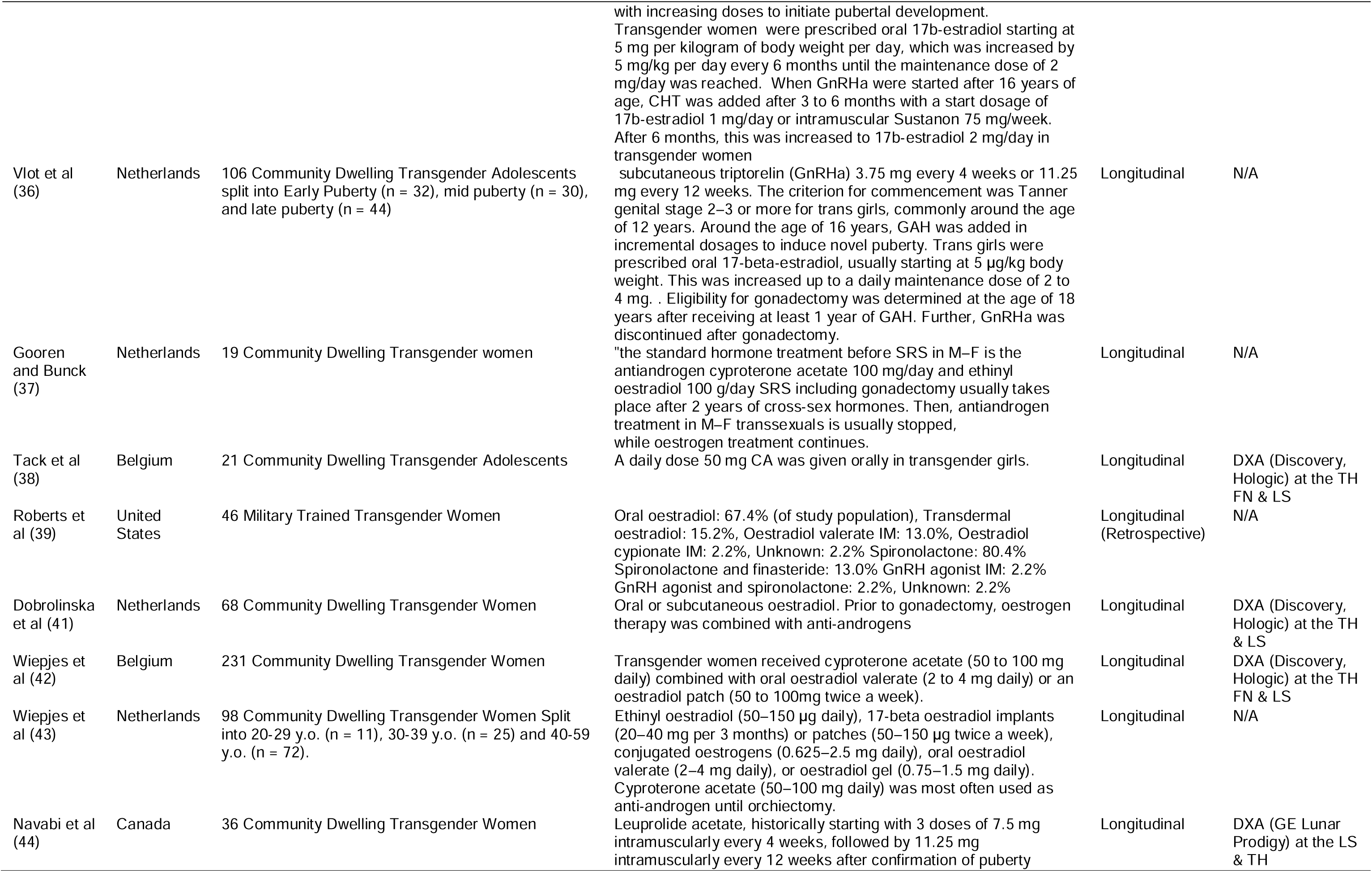
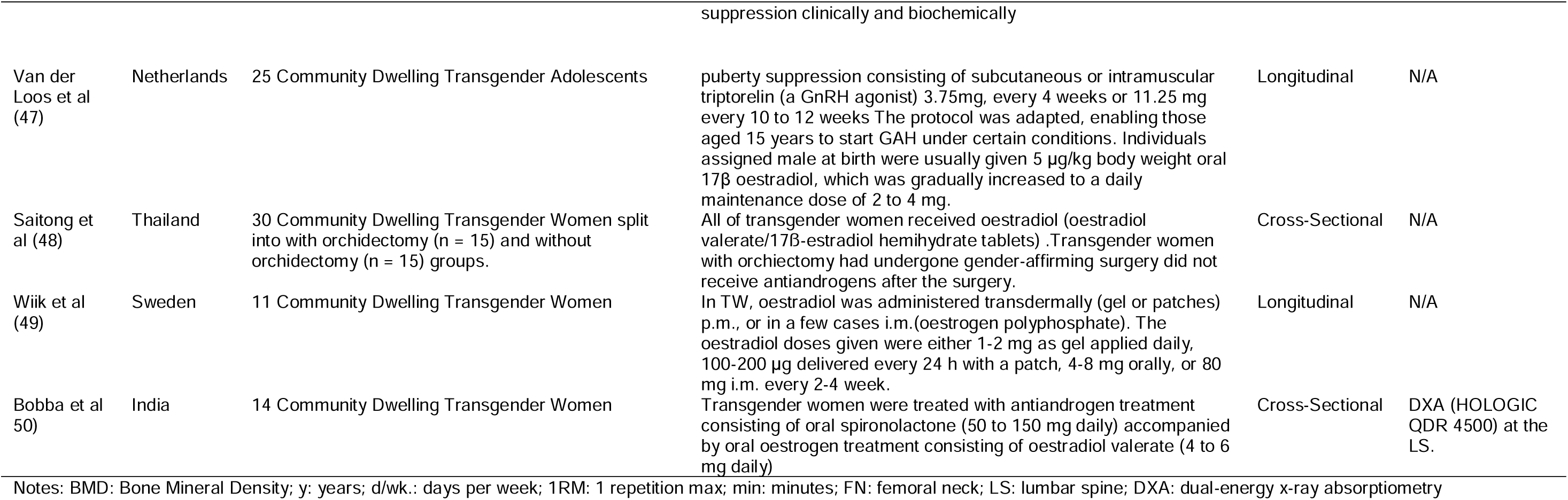
General characteristics of the studies included (*n* = 36)

Of the twenty-eight recent studies included, all were published in peer-reviewed English-language journals, starting in 2016, and ending in 2024. Two studies [52, 53] published before 2016 were included as they were identified during cross-referencing to include outcomes for muscle and body composition that were in the inclusion criteria and not part of the original meta-analysis by Singh-Ospina [21]. The studies were conducted in a variety of countries; twelve in the Netherlands, six in the United States, two in Belgium, Sweden, Brazil, United Kingdom, and Germany and one each in Australia, Spain, Thailand, Italy, South Korea, Italy, Canada, India, Japan and the Czech Republic. The maximum number of Transgender women for which musculoskeletal health assessment was available ranged from 5 to 711 in the intervention groups (mean ± SD= 55 ± 99, [median = 21]) and from 8 to 50 in the comparison groups (mean ± SD = 26 ± 11, [median =24]). Five studies provided sample size estimates [54–58], while the rest did not. The agreement rate on data extraction was 1.0, with no discrepancies between the authors. Two studies [49, 57] did not include all data within the manuscript and following three emails to the corresponding author, there was no response.

### Participant Characteristics

A description of the baseline characteristics of participants can be seen in **Tables 2** and **3**. Reported dropout rates ranged from 0 to 99.9% in the transgender women groups (mean ± SD = 35.5 ± 42.1%, median 22.6%), and there were 0% dropouts in the comparison groups. Fifty-eight per cent of cross-sectional comparisons of transgender women to cisgender men were matched (age and BMI) compared to 42% unmatched.

### GAHT Intervention Characteristics

A description of the GAHT interventions can be seen in **Table 2**. The GAHT interventions varied in length from 1 to 26 years (mean ± SD = 5 ± 6, median 1 year). Compliance with GAHT was not measured in any study. Twelve groups accounted for physical activity, sixteen groups consumed alcohol, and twenty-four groups included smokers. One group contained patients with osteoporosis, and one group included patients using glucocorticoids. No groups presented with osteoarthritis or sarcopenia.

### BMD Assessment Characteristics

All of the studies assessing FN, TH or LS BMD did so using DXA, Fourteen studies used a Hologic DXA system (QDR 4500 Elite [22, 47, 59], Discovery [24, 50, 55, 58, 60–64], horizon [65], or 49-159 [66]), while five studies used a Lunar DXA system [67–71]. One study did not report the DXA model [72]. Insufficient data were reported for the site-specific reliability of the instruments used to assess BMD at either the LS or FN. One study utilised a pQCT device (XCT-2000 [60])

### Muscle Assessment Characteristics

Two studies measured muscle Cross-sectional Area (mCSA), where one utilised a pQCT device (XCT-2000 [60], and two used Magnetic Resonance Imaging [59, 73]. Two studies measured muscular power, with one utilising a JUM001 Jump Mat [74] and the other using both a 400 Series Force Plate and a Monark Peak Bike 894e [75]. Five studies measured muscular strength via handgrip [24, 54, 56, 60, 67], two studies utilised Isokinetic dynamometry [76, 77] and two utilised military fitness tests for strength [78, 79].

### Risk of Bias Assessment

Assessment using the ROBINS-I [36] is shown in **Figure 2** and the **Supplementary Material file RoB** [46]. As can be seen, 39% of the studies were at a low risk of bias, 24% at a moderate risk of bias, 4% at a serious risk of bias and 33% at a critical risk of bias. Given the inability to blind participants in GAHT intervention trials, all studies were at critical risk of bias for the category “Blinding of participants.” The overall risk of bias across all categories was categorised as moderate.

**Figure 2.**
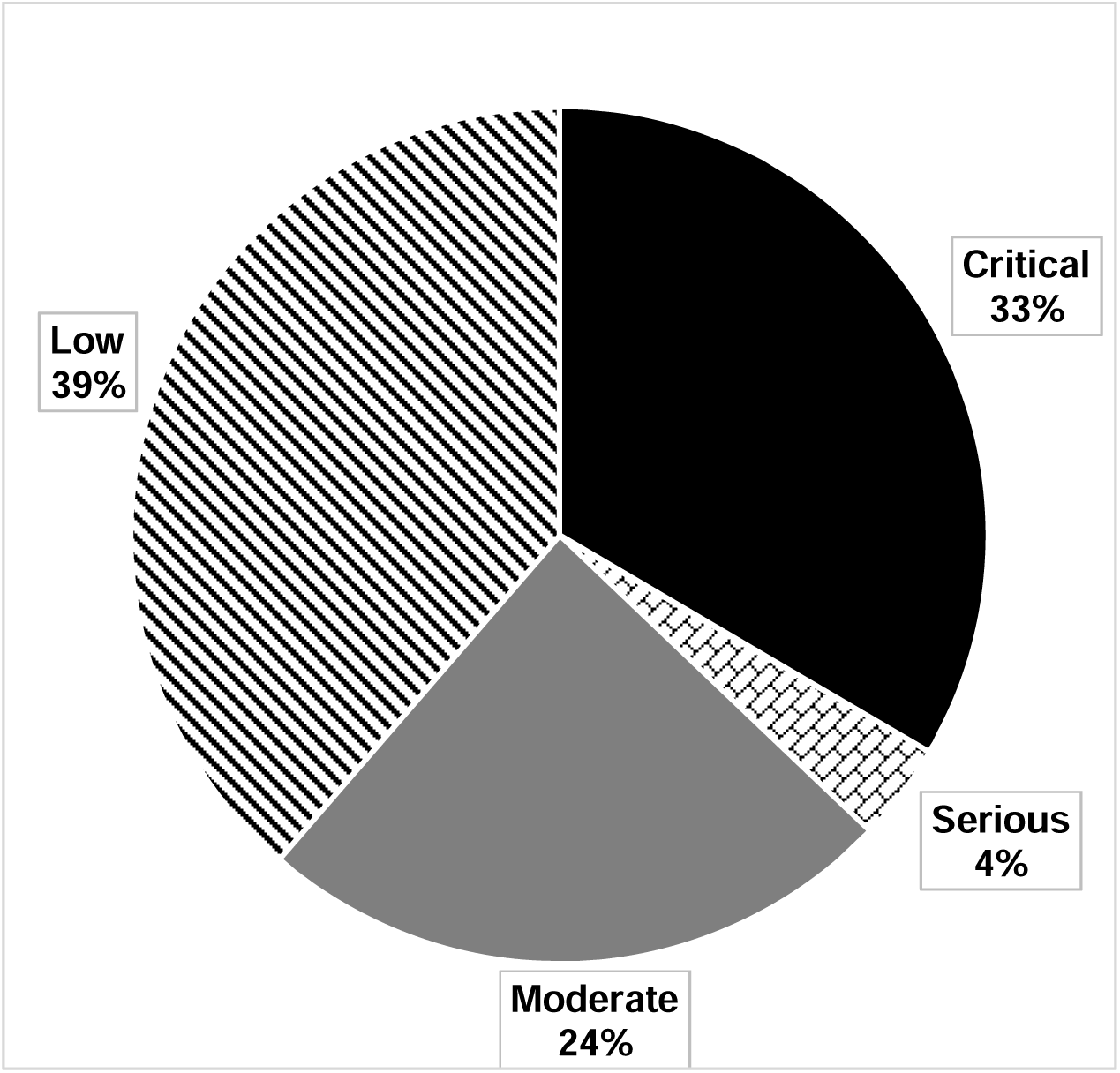
Risk Of Bias In Non-Randomized Studies - of Interventions (ROBINS-I)

### Primary Outcome Measures

#### FN BMD

Longitudinal Changes in FN BMD can be seen in **Table 4** and **Figure 3a**. Longitudinally, a significant increase of FN BMD was found with GAHT (*g* = 0.13 [0.05, 0.20], Z = 3.20, p = 0.00), with no asymmetry (LFK index -0.39, **Supplementary figure 1a**) observed. In addition, no statistically significant heterogeneity was observed (Q = 4.19, p = 0.90), and inconsistency was considered very low (*I^2^* = 0%). Findings were similar when results were collapsed so that only one effect size represented each study. Influence analysis showed that the removal of the Wiepjes et al (2018) group had the biggest influence, with a non-significant effect observed when this study was removed from the analysis (*g* = 0.11 [-0.01, 0.23], Z = 1.82, p = 0.07). The removal of other studies from the analysis did not change the outcome (**Supplementary Table 2**). An evidence profile for changes in FN BMD is shown in online **Supplementary Table 1**. Based on GRADE, the evidence was considered moderate, with future additional studies potentially influencing the overall direction of findings. Cross-sectional comparisons are seen in **Table 4** and **Figure 3b**. Transgender women compared with cisgender men showed no observed statistically significant difference in FN BMD (g = - 0.05 [-1.29, 1.18], Z = -0.08, p = 0.93), with no asymmetry (LFK index = 0.62, **Supplementary figure 1b**) observed.

**Figure 3.**
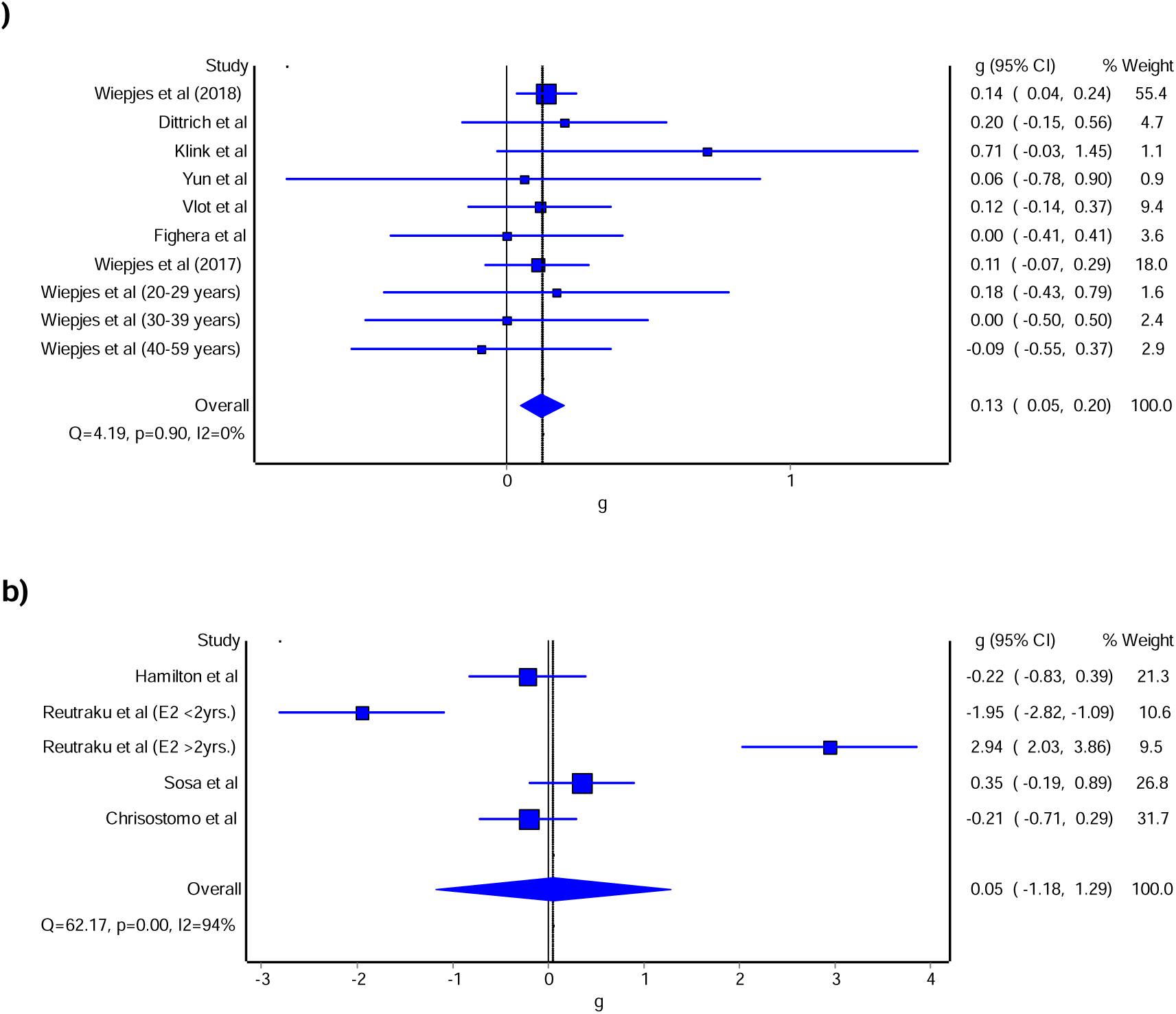
Forest plot for longitudinal (a)changes and cross-sectional comparisons (b) of FN BMD. Forest plot for point estimate standardised effect size changes (g) in FN BMD. The blue squares represent the standardised mean difference (g) while the left and right extremes of the squares represent the corresponding 95% confidence intervals. The middle of the blue diamond represents the overall standardised mean difference (g) while the left and right extremes of the diamond represent the corresponding 95% confidence intervals.

**Table 3.** Baseline characteristics of participants

| Variable | Transgender Women |  |  |  |  | Cisgender Male Comparators |  |  |  |  |
| --- | --- | --- | --- | --- | --- | --- | --- | --- | --- | --- |
|  | Groups (#) | Participants (#) | Mean ± SD | Mdn | Range | Groups (#) | Participants (#) | Mean ± SD | Mdn | Range |
| Age (y) | 38 | 2261 | 30 ± 5 | 22 | 21 - 43 | 9 | 284 | 32 ± 7 | 29 | 24 - 44 |
| BMI (kg/m<sup>2</sup>) | 15 | 645 | 23.1 ± 1.7 | 23.1 | 20.3 – 26.1 | 6 | 123 | 24.1 ± 1.9 | 23.3 | 22.5 – 26.7 |
| Fat Mass (kg) | 10 | 242 | 15.9 ± 2.7 | 15.1 | 12.7 – 22.1 | 2 | 33 | 17.8 ± 3.4 | 17.8 | 15.4 - 20.2 |
| Fat-Free Mass (kg) | 12 | 242 | 35.1 ± 20.8 | 33.8 | 5.6 - 62.6 | 6 | 128 | 55.4 ± 18.9 | 56.1 | 20.5 – 72.1 |
| BMD (g/cm<sup>2</sup>) |  |  |  |  |  |  |  |  |  |  |
| Femoral Neck | 7 | 1195 | 0.80 ± 0.12 | 0.80 | 0.79 - 1.07 | 5 | 124 | 0.98 ± 0.10 | 1.01 | 0.83 - 1.11 |
| Lumbar Spine | 13 | 1376 | 1.03 ± 0.10 | 1.00 | 0.84 - 1.20 | 5 | 124 | 1.12 ± 0.08 | 1.14 | 1.03 - 1.23 |
| Total Hip | 5 | 1174 | 0.96 ± 0.05 | 0.94 | 0.93 - 1.04 | 4 | 98 | 1.08 ± 0.02 | 1.07 | 1.07 - 1.11 |

Notes: #, Number of; SD, Standard Deviation; Mdn, Median; y, years; BMI, Body Mass Index; BMD, Bone Mineral Density; g/cm, grams per centimetre.

**Table 4.**
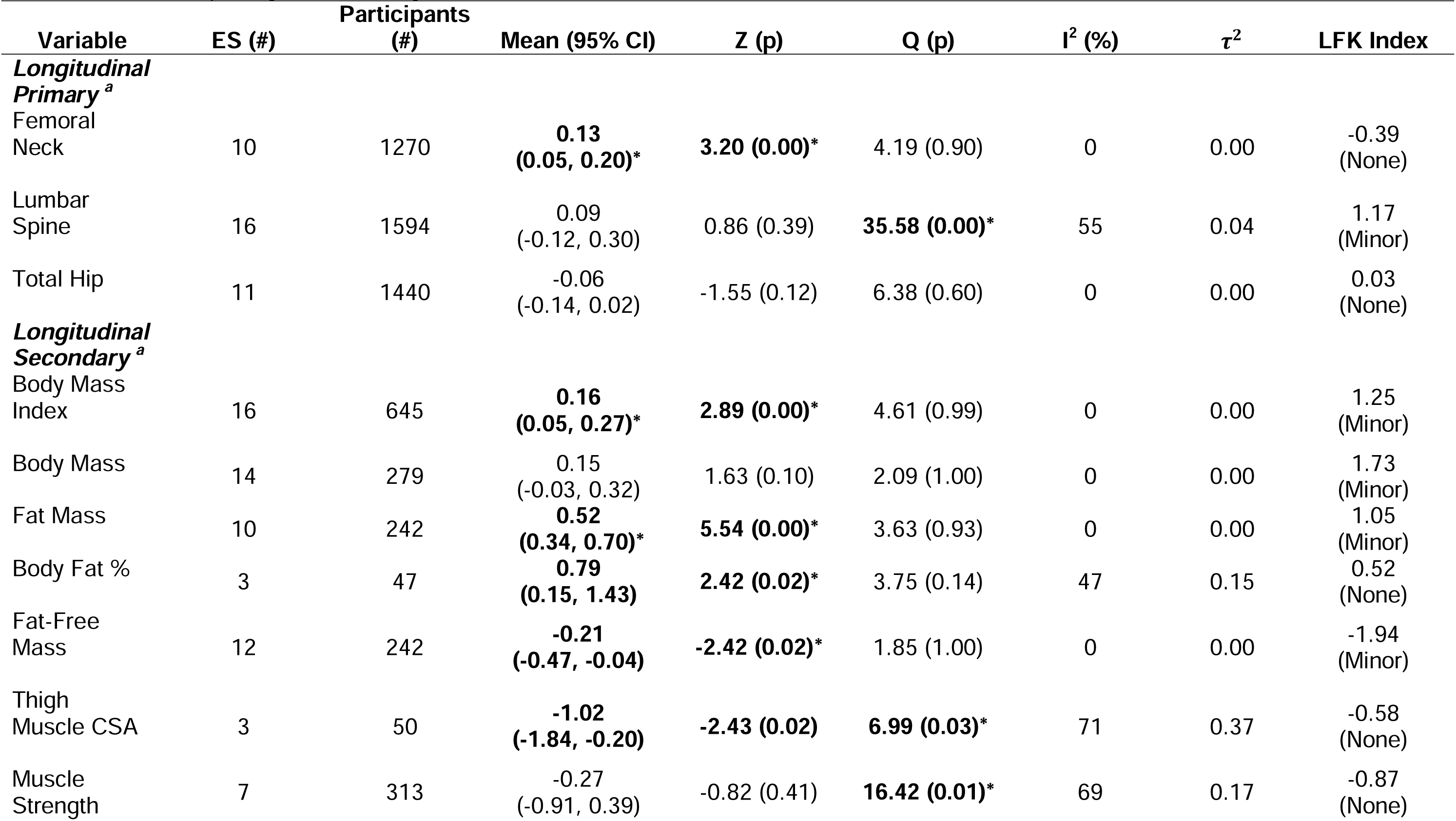
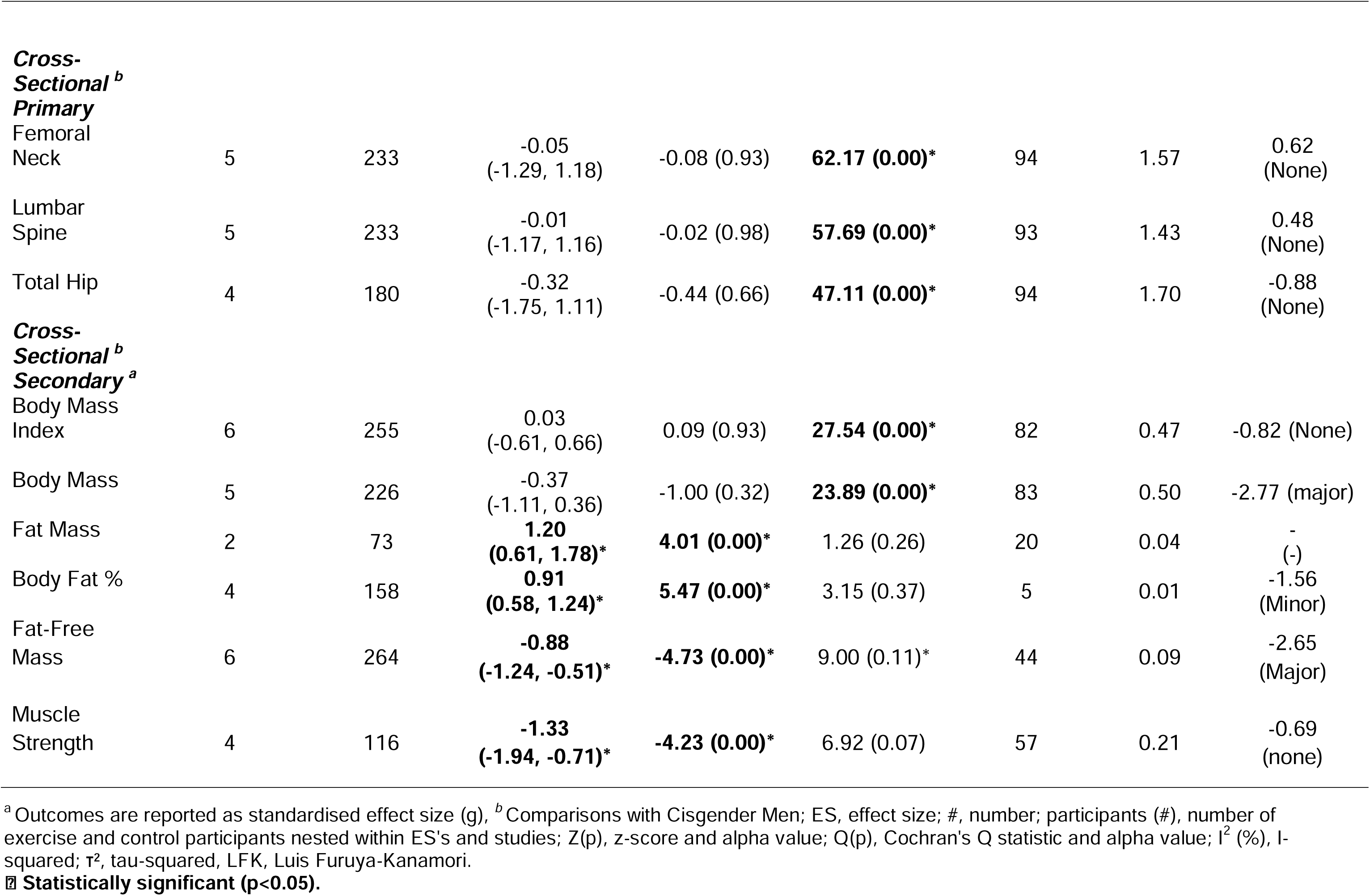
Results of primary and secondary outcomes

#### LS BMD

Longitudinal changes in LS BMD can be seen in **Table 4** and **Figure 4a**. There was no observed statistically significant change with GAHT on LS BMD (g = 0.09 [-0.12, 0.30], Z = 0.86, p = 0.39), with minor asymmetry (LFK index 1.17, **Supplementary figure 2a**) observed. In addition, statistically significant heterogeneity was observed (Q = 35.58, p = 0.00), and inconsistency was considered moderate (*I^2^*= 55%). Findings were similar when results were collapsed so that only one effect size represented each study. With the removal of each effect size from the model, the results remained non-significant. An evidence profile for changes in LS BMD is shown in online **supplementary table 1**. Based on GRADE, the evidence was considered low, with future additional studies likely to influence the overall direction of findings. Transgender women compared with cisgender men cross-sectionally showed no observed statistically significant difference in LS BMD (g = -0.01 [-1.17, 1.16], Z = -0.02, p = 0.98), with no asymmetry (LFK index = -0.50, **Supplementary figure 2b**) observed.

**Figure 4.**
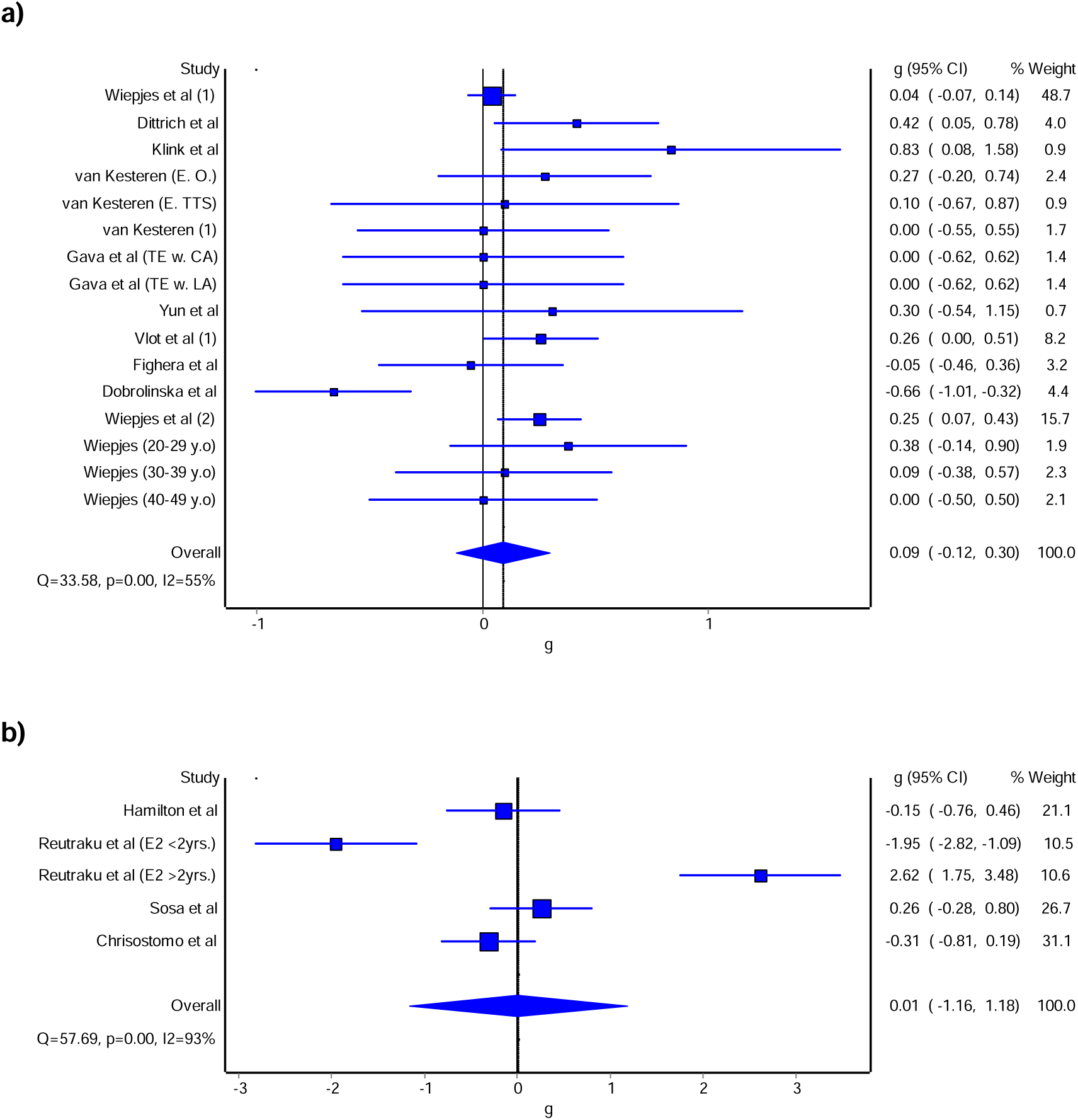
Forest plot for longitudinal changes with GAHT in transgender women a) and cross-sectional comparisons with cisgender men b) of LS BMD. The blue squares represent the standardised mean difference (g) while the left and right extremes of the squares represent the corresponding 95% confidence intervals. The middle of the blue diamond represents the overall standardised mean difference (g) while the left and right extremes of the diamond represent the corresponding 95% confidence intervals.

#### TH BMD

Longitudinal changes in TH BMD can be seen in **Table 4** and **Figure 5a**. There was no observed statistically significant change with GAHT on TH BMD (g = -0.06 [-0.14, 0.02], Z = - 1.55, p = 0.12), with no asymmetry (LFK index = 0.03, **Supplementary figure 3a**) observed. In addition, no statistically significant heterogeneity was observed (Q = 6.38, p = 0.60), and inconsistency was considered very low (*I^2^* = 0%). The results remained non-significant after removing each effect size from the model. An evidence profile for changes in TH BMD is shown in online **supplementary table 1**. Based on GRADE, the evidence was considered moderate, with future additional studies potentially influencing the overall direction of findings. Transgender women compared with cisgender men cross-sectionally showed no observed statistically significant difference in TH BMD (g = -0.32 [-1.75, 1.11], Z = -0.44, p = 0.66), with no asymmetry (LFK index = -0.88, **Supplementary figure 3b**) observed. An evidence profile for changes in TH BMD is shown in online **Supplementary Table 1.**

**Figure 5.**
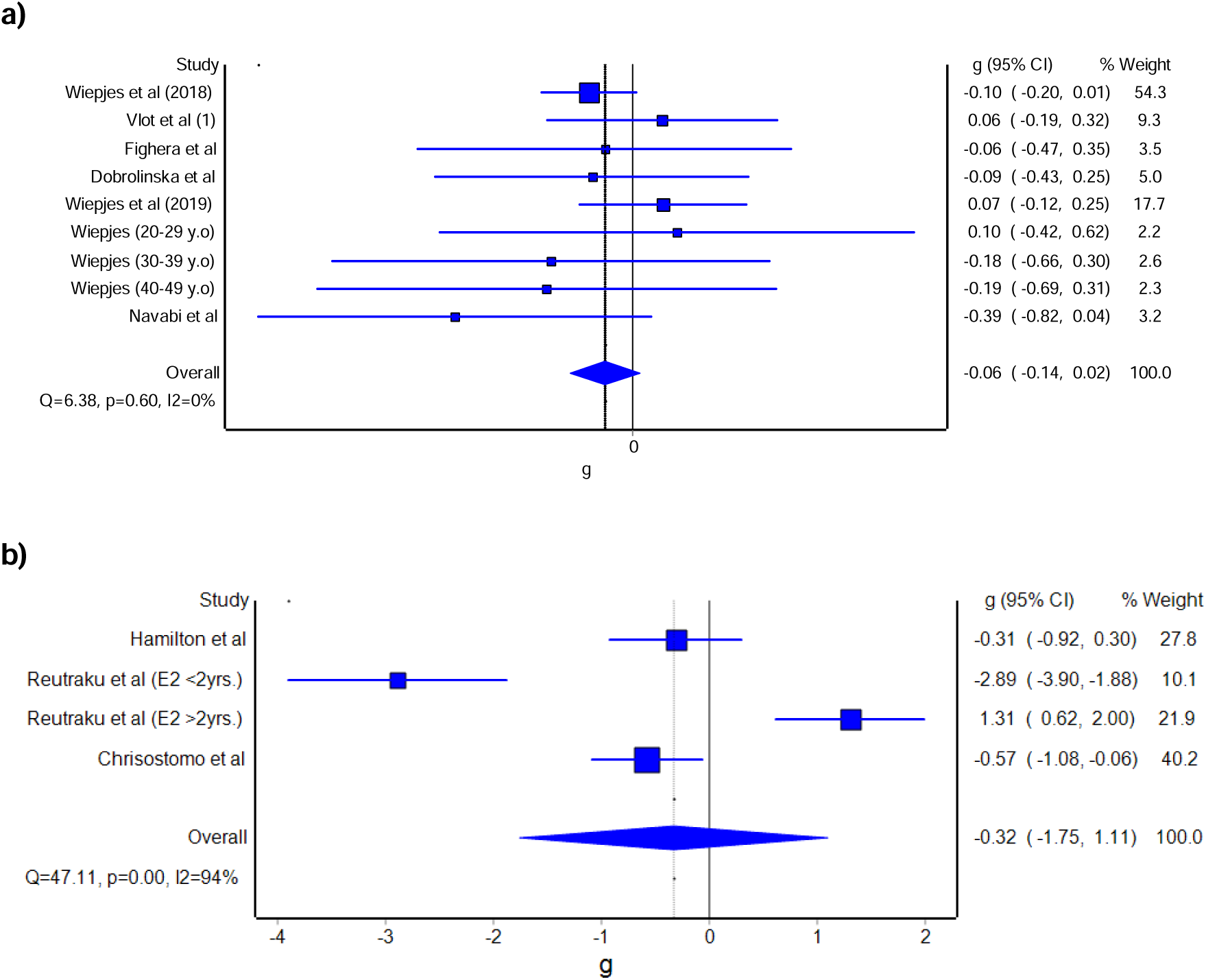
Forest plot for longitudinal changes with GAHT in transgender women a) and cross-sectional comparisons with cisgender men b) of TH BMD. Forest plot for point estimate standardised effect size changes (g) in TH BMD. The blue squares represent the standardised mean difference (g) while the left and right extremes of the squares represent the corresponding 95% confidence intervals. The middle of the blue diamond represents the overall standardised mean difference (g) while the left and right extremes of the diamond represent the corresponding 95% confidence intervals.

### Changes in Secondary Outcomes

#### Body Mass Index (BMI)

Longitudinal changes in BMI in transgender women with GAHT can be seen in **Table 4** and **Figure 6a**. There was an observed statistically significant effect of GAHT on BMI (g = 0.16 [0.05, 0.27], Z = 2.89, p = 0.00), with minor asymmetry (LFK index = 1.25), **Supplementary** Figure 4a) observed. In addition, no statistically significant heterogeneity was observed (Q = 4.61, p = 0.99), and overall inconsistency was considered to be low (*I^2^* = 0%). Influence analysis showed that the removal of other studies from the analysis did not change the outcome. Based on GRADE, the evidence was considered moderate, with future additional studies potentially influencing the overall direction of findings. Transgender women compared with cisgender men cross-sectionally showed no observed statistically significant difference in BMI (g = 0.03 [-0.61, 0.66], Z = 0.09, p = 0.93, **Figure 6b**), with no asymmetry (LFK index = -0.82, **Supplementary figure 4b**) observed. An evidence profile for changes in BMI is shown in online **Supplementary Table 1.**

**Figure 6.**
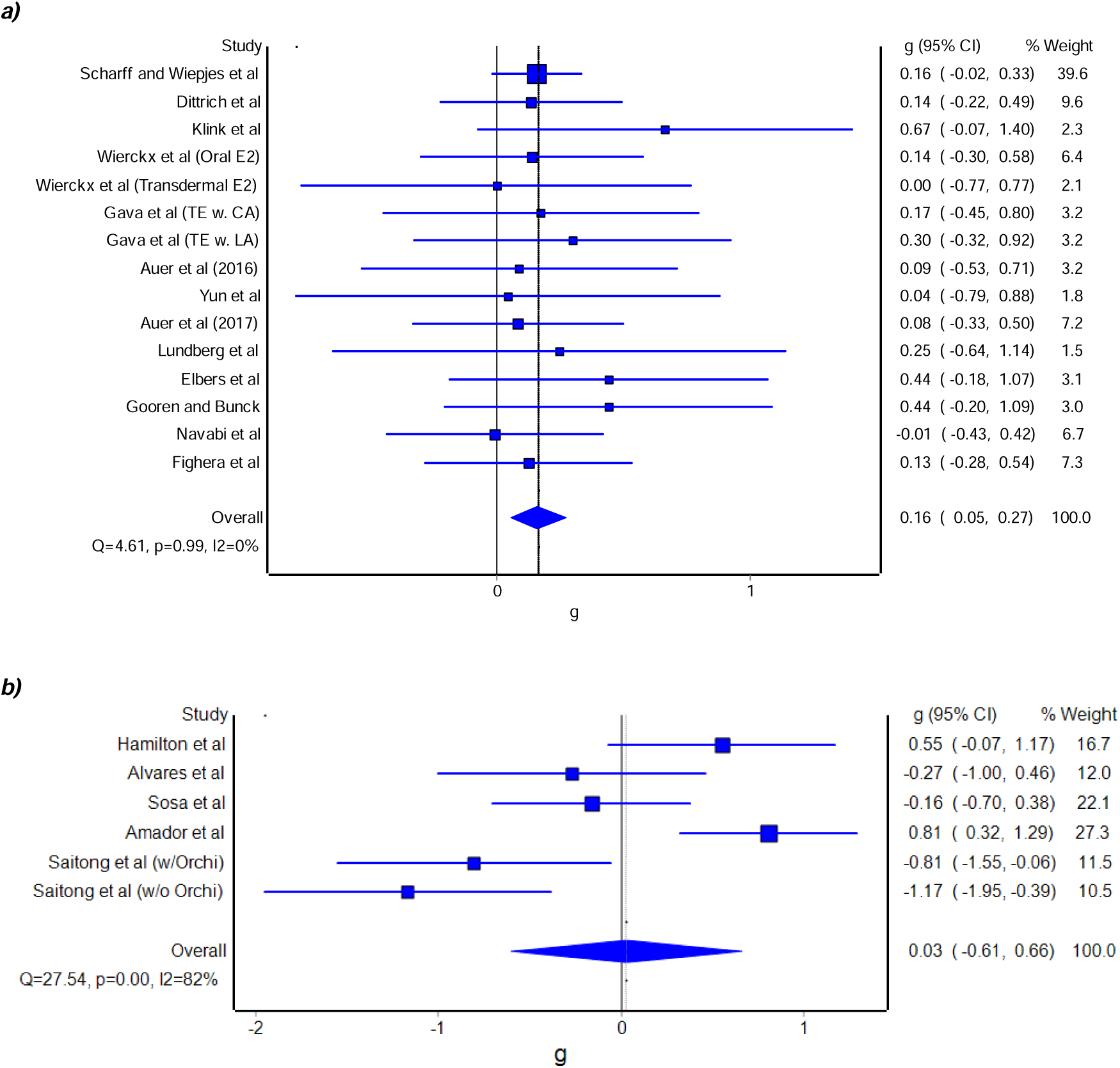
Forest plot for longitudinal changes with GAHT in transgender women a) and cross-sectional comparisons with cisgender men b) of BMI. Forest plot for point estimate standardised effect size changes (g) BMI. The blue squares represent the standardised mean difference (g) while the left and right extremes of the squares represent the corresponding 95% confidence intervals. The middle of the blue diamond represents the overall standardised mean difference (g) while the left and right extremes of the diamond represent the corresponding 95% confidence intervals.

#### Body Mass (BM)

Longitudinal changes of BM in transgender women with GAHT can be seen in **Table 4** and **Figure 7a**. There was no statistically significant effect of GAHT on BM (g = 0.15 [-0.03, 0.32], Z = 1.63, p = 0.10), with minor asymmetry (LFK index = 1.73), **Supplementary** Figure 5a) observed. In addition, no statistically significant heterogeneity was observed (Q = 2.09, p = 1.00 and overall inconsistency was considered to be low (*I^2^* = 0%). Influence analysis showed that the removal of other studies from the analysis did not change the outcome. Based on GRADE, the evidence was considered moderate, with future additional studies potentially influencing the overall direction of findings. Transgender women compared with cisgender men cross-sectionally showed no observed statistically significant difference in BM (g = -0.37 [-1.11, 0.36], Z = -1.00, p = 0.32, **Figure 7b**), with major asymmetry (LFK index = -2.77, **Supplementary figure 5b**) observed. An evidence profile for changes in BM is shown in online **Supplementary Table 1.**

**Figure 7.**
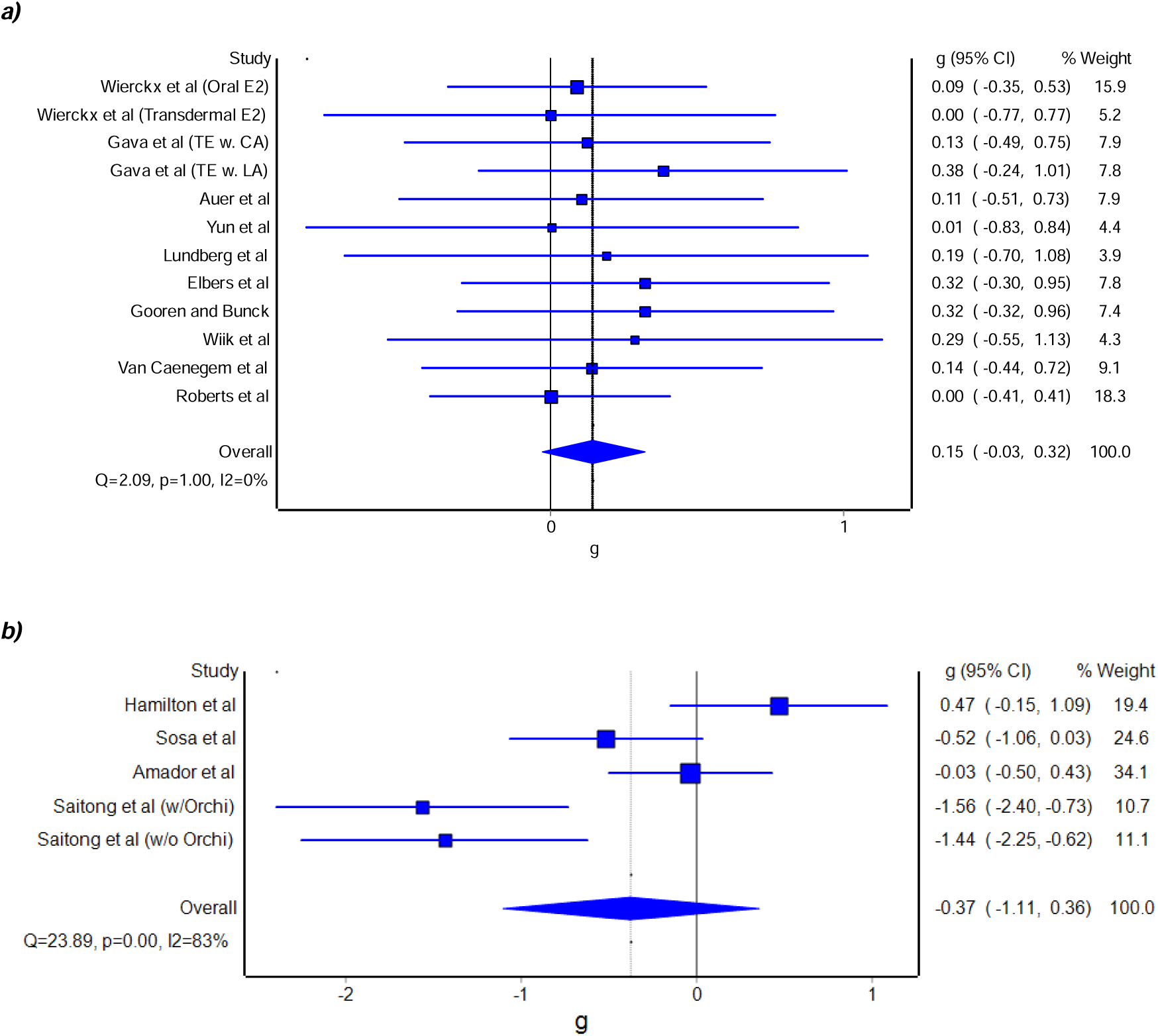
Forest plot for longitudinal changes with GAHT in transgender women a) and cross-sectional comparisons with cisgender men b) of Body Mass. The blue squares represent the standardised mean difference (g) while the left and right extremes of the squares represent the corresponding 95% confidence intervals. The middle of the blue diamond represents the overall standardised mean difference (g) while the left and right extremes of the diamond represent the corresponding 95% confidence intervals.

#### Fat Mass (FM)

Longitudinal Results of FM in transgender women with GAHT can be seen in **Table 4** and **Figure 8a**. There was observed statistically significant effect of GAHT on FM (g = 0.52 [0.34, 0.70], Z = 5.54, p = 0.00), with minor asymmetry (LFK index 1.05), **Supplementary figure 6a**) observed. In addition, no statistically significant heterogeneity was observed (Q = 3.63, p = 0.93), and overall inconsistency was considered to be very low (*I^2^* = 0%). Influence analysis showed that the removal of any study did not change the outcome. Based on GRADE, the evidence was considered high, and further research is unlikely to significantly change the confidence in the estimate. Transgender women compared with cisgender men cross-sectionally showed an observed statistically significant difference in FM (g = 1.20 [0.61, 1.78], Z = 4.01, p = 0.00, **Figure 8b**) with minor asymmetry (LFK index = 1.05, **Supplementary figure 6b**). An evidence profile for changes in FM is shown in online **Supplementary Table 1.**

**Figure 8.**
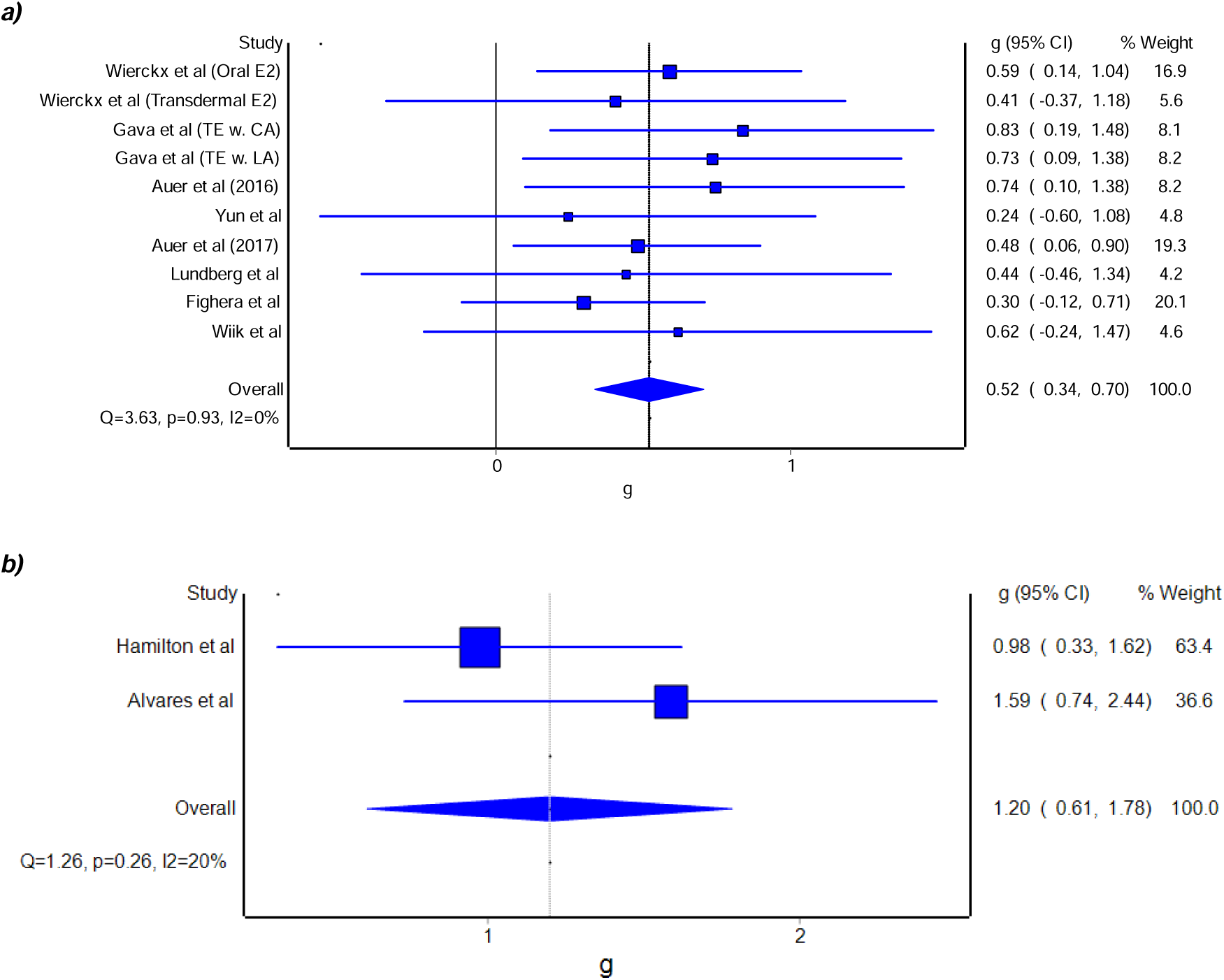
Forest plot for longitudinal changes with GAHT in transgender women a) and cross-sectional comparisons with cisgender men b) of Fat Mass. The blue squares represent the standardised mean difference (g) while the left and right extremes of the squares represent the corresponding 95% confidence intervals. The middle of the blue diamond represents the overall standardised mean difference (g) while the left and right extremes of the diamond represent the corresponding 95% confidence intervals.

#### Body Fat % (BF%)

Longitudinal Results of BF% in transgender women with GAHT can be seen in **Table 4** and **Figure 9a**. There was an observed statistically significant effect of GAHT on BF% (g = 0.79 [0.15, 1.43], Z = 2.42, p = 0.02), with no asymmetry (LFK index 0.52), **Supplementary figure 7a**) observed. In addition, no significant heterogeneity was observed (Q = 3.75, p = 0.14), and overall inconsistency was considered to be low (*I^2^* = 47%). Based on GRADE, the evidence was considered moderate, with future additional studies potentially influencing the overall direction of findings. Transgender women compared with cisgender men cross-sectionally showed an observed statistically significant difference in BF% (g =0.91 [0.58, 1.24], Z = 5.47, p = 0.00, **Figure 9b**), with minor asymmetry (LFK index = -1.56, **Supplementary figure 7b**) observed. An evidence profile for changes in BF% is shown in online **Supplementary Table 1.**

**Figure 9.**
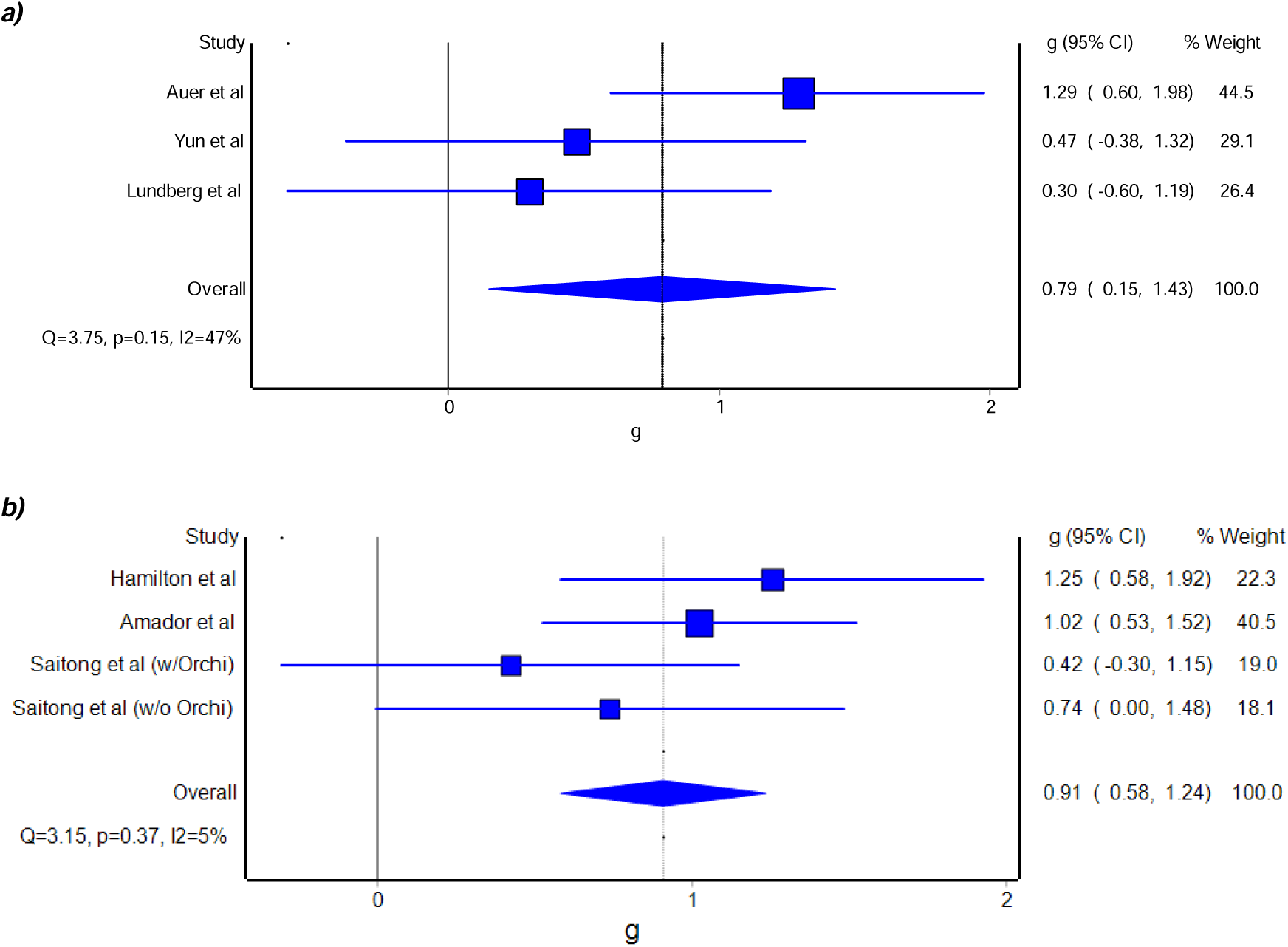
Forest plot for longitudinal changes with GAHT in transgender women a) and cross-sectional comparisons with cisgender men b) of Body Fat %. The blue squares represent the standardised mean difference (g) while the left and right extremes of the squares represent the corresponding 95% confidence intervals. The middle of the blue diamond represents the overall standardised mean difference (g) while the left and right extremes of the diamond represent the corresponding 95% confidence intervals.

#### Fat-Free Mass (FFM)

Longitudinal Results of FFM in transgender women with GAHT can be seen in **Table 4** and **Figure 10a**. There was an observed statistically significant effect of GAHT on FFM (g = - 0.21 [-0.47, -0.04, Z = -2.42, p = 0.02), with minor asymmetry (LFK index = -1.94, **Supplementary figure 8a**) observed. In addition, no heterogeneity was observed (Q = 1.85, p = 1.00), and overall inconsistency was considered to be very low (*I^2^* = 0%). Influence analysis showed that the removal of any study did not change the outcome (**Supplementary Table 1**). Based on GRADE, the evidence was considered moderate, with future additional studies potentially influencing the overall direction of findings. Transgender women compared with cisgender men cross-sectionally showed an observed statistically significant difference in FFM Mass (g = -0.88 [-1.24, -0.51], Z = -4.73, p = 0.00, **Figure 10b**), with minor asymmetry (LFK index = -2.65, **Supplementary figure 8b**) observed. An evidence profile for changes in FFM is shown in online **Supplementary Table 1.**

**Figure 10.**
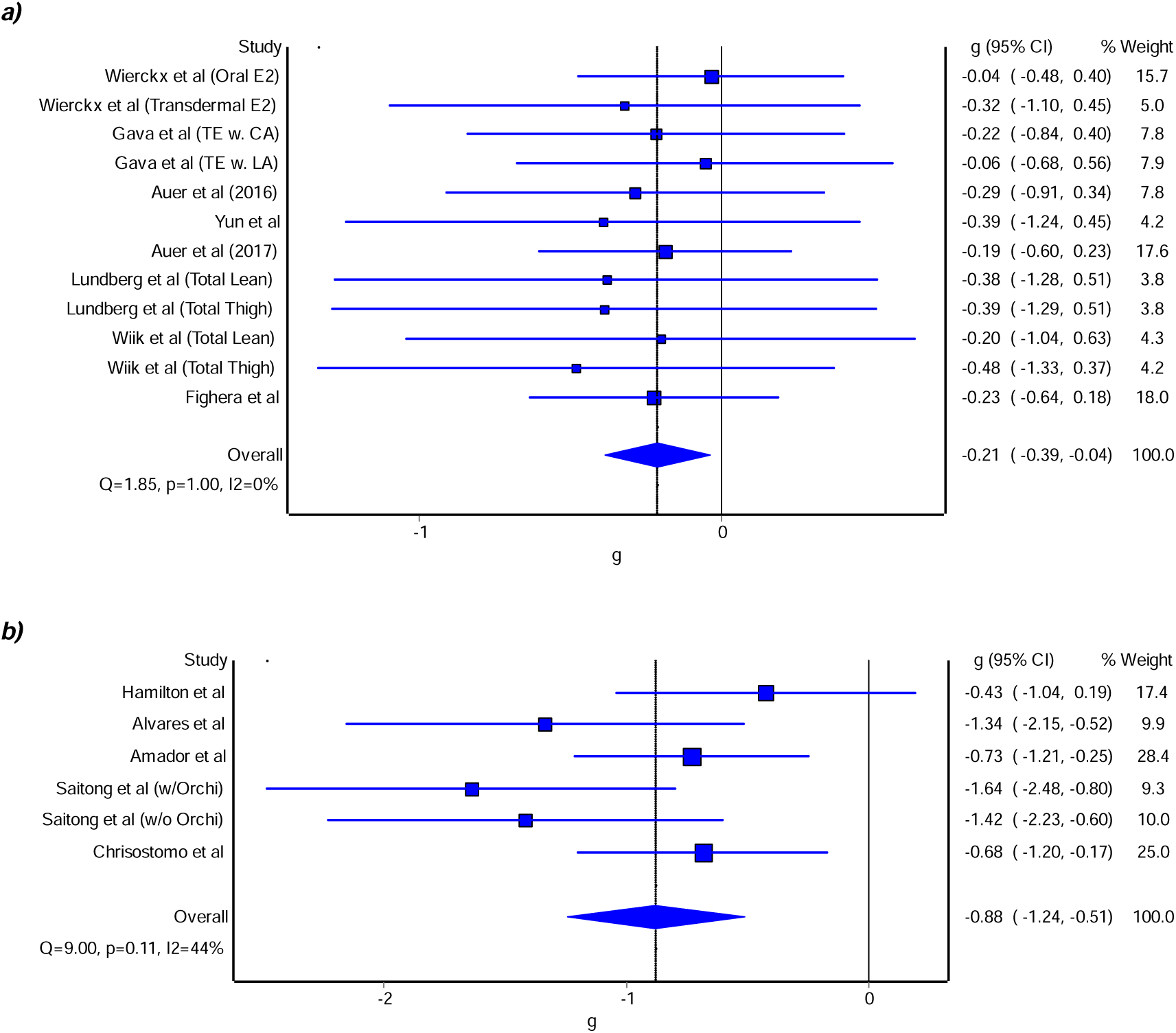
Forest plot for longitudinal changes with GAHT in transgender women a) and cross-sectional comparisons with cisgender men b) of Fat Free Mass. The blue squares represent the standardised mean difference (g) while the left and right extremes of the squares represent the corresponding 95% confidence intervals. The middle of the blue diamond represents the overall standardised mean difference (g) while the left and right extremes of the diamond represent the corresponding 95% confidence intervals.

#### Muscle Cross-Sectional Area (mCSA)

Longitudinal Results of mCSA in transgender women with GAHT can be seen in **Table 4** and **Figure 11**. There was an observed statistically significant effect of GAHT on MCSA (g = - 1.02 [-1.84, -0.20] Z = -2.43 p = 0.02), with no asymmetry (LFK index= -0.58, **Supplementary figure 9a**) observed. In addition, statistically significant heterogeneity was observed (Q = 19.13, p = 0.00), and overall inconsistency was considered to be moderate (*I^2^* = 69%). Influence analysis showed that the removal of the Elbers et al group had the biggest influence, with no significant effect observed when this study was removed from the analysis (*g* = -1.17 [-2.62, 0.27], Z = -1.59, p = 0.11). (**Supplementary Table 3**). Based on GRADE, the evidence was considered moderate, with future additional studies potentially influencing the overall direction of findings. There was no data to compare Transgender women with cisgender men cross-sectionally for mCSA. An evidence profile for changes in mCSA is shown in online **Supplementary Table 1.**

**Figure 11.**
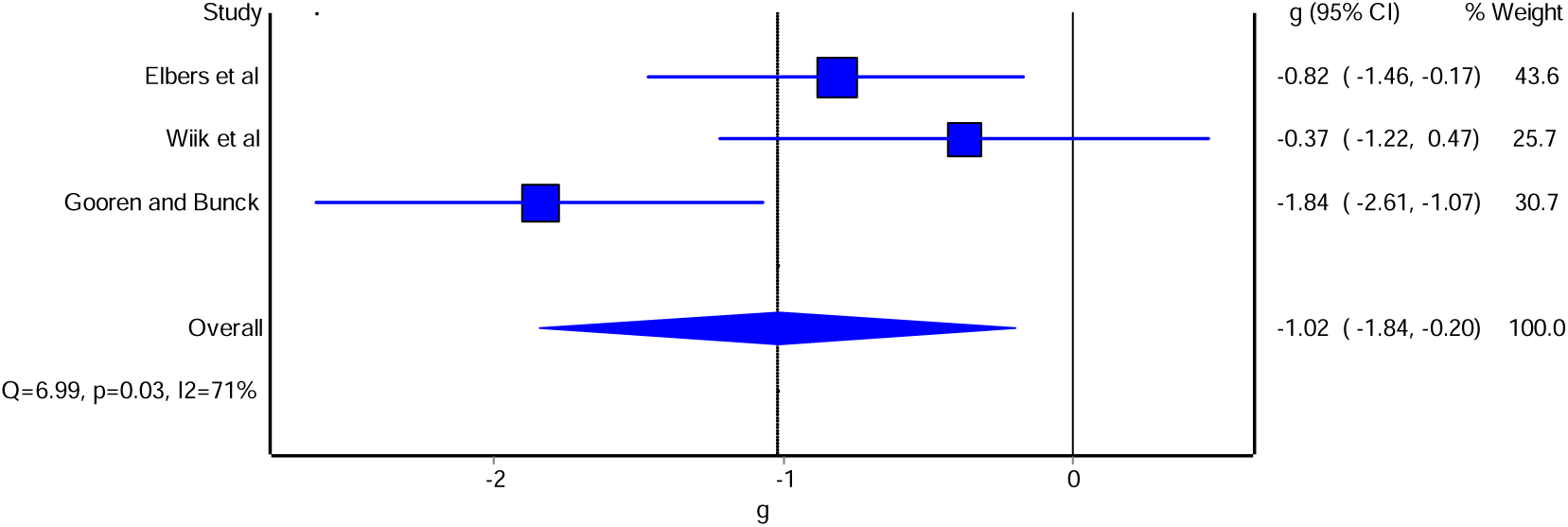
Forest plot for longitudinal changes with GAHT in transgender women’s Thigh Muscle Cross Sectional Area. The blue squares represent the standardised mean difference (g) while the left and right extremes of the squares represent the corresponding 95% confidence intervals. The middle of the blue diamond represents the overall standardised mean difference (g) while the left and right extremes of the diamond represent the corresponding 95% confidence intervals.

#### Muscle Strength

Longitudinal Results of Muscle Strength in transgender women with GAHT can be seen in **Table 4** and **Figure 12a**. There was no observed statistically significant effect of GAHT on Muscle Strength (g = -0.27 [-0.91, 0.39], Z = -0.82, p = 0.41), with no asymmetry (LFK index = -0.87), **Supplementary figure 10a**) observed. In addition, statistically significant heterogeneity was observed (Q = 55.52, p = 0.00), and overall inconsistency was considered to be moderate (*I^2^* = 75%). Influence analysis showed that when removed, the Chiccarelli et al group had the biggest influence on the outcome, and the outcome became a significant effect (*g* = -0.17 [-0.33, -0.02], Z = -2.17, p = 0.03, **Supplementary Table 4**). Based on GRADE, the evidence was considered low, with future additional studies likely to influence the overall direction of findings. Transgender women compared with cisgender men cross-sectionally showed an observed statistically significant difference in Muscle Strength (g = - 1.69 [-2.06, -1.38], Z = -8.86, p = 0.00, **Figure 12b**), with no asymmetry (LFK index = -0.87, **Supplementary figure 10b**) observed. An evidence profile for changes in Muscle Strength is shown in online **Supplementary Table 1.**

**Figure 12.**
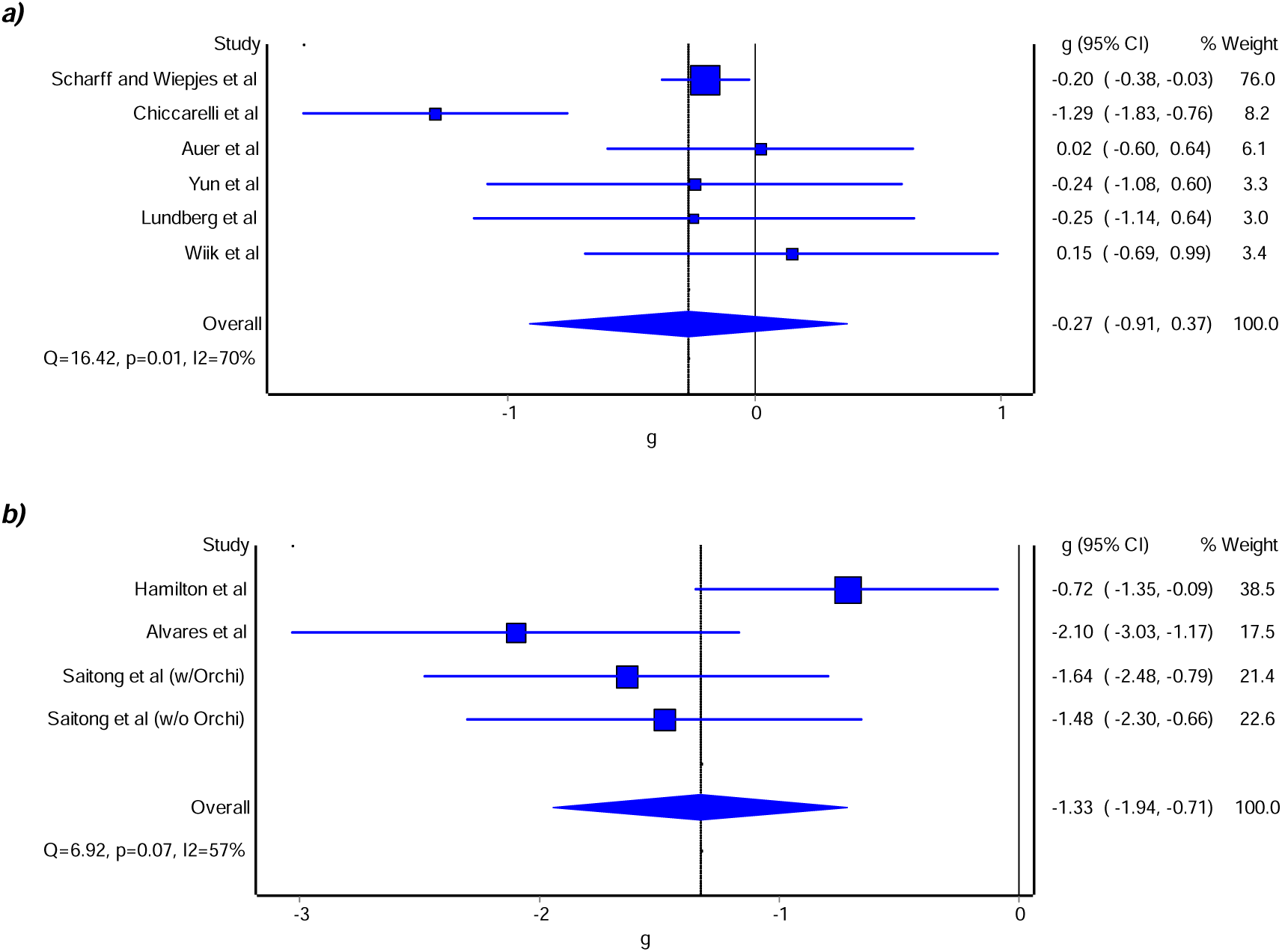
Forest plot for longitudinal changes with GAHT in transgender women a) and cross-sectional comparisons with cisgender men b) of Muscle Strength. The blue squares represent the standardised mean difference (g) while the left and right extremes of the squares represent the corresponding 95% confidence intervals. The middle of the blue diamond represents the overall standardised mean difference (g) while the left and right extremes of the diamond represent the corresponding 95% confidence intervals.

## Discussion

The primary aim of the current systematic review and meta-analysis was to update the work by Singh-Ospina [21] examining the effects of GAHT on the Bone Health of Transgender Women. The overall findings suggest that longitudinal GAHT is associated with a statistically significant benefit in FN but not TH or LS BMD. The beneficial effect observed in FN BMD is larger and statistically significant, compared to the previously shown no effect by Singh- Ospina [21]. However, due to the greater number of effect sizes included, the use of a more robust meta-analytical methodology, and the low heterogeneity and inconsistency (**Table 4**), the observed effect in this meta-analysis provides moderate evidence for the beneficial effect of GAHT on FN BMD in transgender women. As peak bone mass in individuals assigned male at birth is typically reached by approximately 27 years of age [80], and the transgender women in this study had a mean age of 30 ± 5 years (Table 3), and the influence of age-related bone gains is expected to be minimal. This strengthens the validity of the findings presented. Based on GRADE, the available evidence is sufficient to support a conclusion, but further research may still impact the confidence in the estimate [81]**(Supplementary Table 1).**

While the current study provides secure evidence of a beneficial effect of longitudinal GAHT on FN BMD, it does not strengthen the evidence of a positive longitudinal effect in LS BMD. Singh-Ospina [21] and Fighera [82] describe a positive effect of GAHT on LS BMD. The pooled effect size after 24 months of GAHT in Singh-Ospina [21] was 175% higher than was observed in this updated analysis after an average of 5 years of GAHT, while the effect size in Fighera [82] was 77% below yet significant after 12 months and not significant after 24 months. The greater number of studies included and the more robust methods in this analysis should normally suggest that this effect is unlikely to be influenced by additional research. However, our longitudinal LS BMD results are plagued with significant heterogeneity and moderate inconsistency, indicating that the studies may not be comparable enough to be pooled together and interpreted as a single, unified effect size. Together with the low-GRADE outcome, further work on the longitudinal effects of GAHT on LS BMD is recommended. Concurring with Singh-Ospina [21] original analysis and Fighera [82]TH BMD seems to remain unaffected by longitudinal GAHT, as our results showed no statistically significant effect with very low heterogeneity and inconsistency found in the analysis. For TH, the available evidence is sufficient to support a conclusion, but further research may still impact the confidence in the estimate (Supplementary Table 2)[81].

The findings for FN BMD were sensitive to influence analysis, where the effect remained significant on the removal of all effect sizes except for the Wiepjes et al (2018) group (**Supplementary Table 2**). When this group was removed from the model, the overall effect became non-significant, with the 95% confidence interval for the mean effect crossing zero. The opposite was true for LS and TH BMD, which remained non-significant through influence analysis. To the best of our knowledge, Singh-Ospina [21] did not perform an influence analysis for BMD outcomes, therefore, this strengthens our updated analysis’s conclusions.

Our secondary aim was to update the work of Singh-Ospina [21] and extend the analysis to overall musculoskeletal health, bringing insights into longitudinal GAHT effects. The overall findings suggest that longitudinal GAHT is associated with statistically significant gains in BMI, FM, BF% and BM (Table 4). GAHT was also associated with detrimental effects on FFM and MCSA (Table 4). GAHT was not associated with declines in Muscle Strength (Table 4) although this was sensitive to influence analysis where the effect became a significant decline (**Supplementary Table 4**).

In cross-sectional comparisons with Cisgender men, Transgender women showed higher FM and BF%, while showing less FFM and muscle strength. However, FFM and Muscle Strength presented with significant heterogeneity and high inconsistency, suggesting variability across studies, potentially due to differences in study design, participant characteristics (e.g., age at GAHT initiation, duration of treatment, or physical activity levels), or methodological approaches to the assessment. This variability limits the ability to draw definitive conclusions about the comparisons of FFM in transgender women relative to cisgender men and highlights the need for further research with more standardised methodologies and well-matched comparator groups. Given that FM, and BF%, do not exhibit the same level of heterogeneity and inconsistency as fat-free mass and Muscle Strength, these findings can be interpreted with greater confidence. The consistent finding of higher fat mass and body fat % in transgender women compared to cisgender men suggests a reliable difference in adipose tissue distribution. However, all cross-sectional measures produced low to very low evidence based on GRADE (**Supplementary Table 1**). Therefore, further cross-sectional comparisons between cisgender men and transgender women are recommended.

Longitudinal studies indicate that GAHT does not reduce muscle strength. However, this was sensitive to influence analysis and had three different modes of testing (Handgrip, Isokinetic Dynamometry and fitness testing), resulting in high heterogeneity, suggesting that more robust studies may find a different outcome. Handgrip use as a measure of over all muscle strength in transgender women is problematic, as the test is dependent upon finger length, finger span, and hand perimeter, which are unaltered with GAHT; therefore, masking the true extent of muscle strength changes [83]. While transgender women exhibit lower muscle strength than cisgender men in cross-sectional analyses, this may reflect pre-existing differences, training status, or muscle quality rather than GAHT-induced declines. This discrepancy underscores the need for caution when interpreting cross-sectional data, especially as only 58% of the cross-sectional data were of matched cohorts. While longitudinal GAHT alters body composition as shown in Table 4, its direct effects on muscle strength may be minimal or counterbalanced by neuromuscular adaptation and physical activity. Further longitudinal research with well-matched comparators is critical to clarify muscle strength trajectories in transgender women undergoing GAHT, given the high heterogeneity and inconsistency shown in the results of muscle strength (**Table 4),** together with the significant results of the influence analysis **(Supplementary Table 4**).

### Limitations

The observed increase in femoral neck BMD (g = 0.13) reflects a small effect size and demonstrates borderline statistical significance when a single influential study is excluded, thereby raising concerns regarding the robustness of this finding. Given the modest magnitude of this effect, it is unlikely to be clinically meaningful in most contexts. Specifically, it does not approach the threshold generally considered indicative of a clinically important change; namely, an observed increase of at least 5.5% (approximately g ≈ 0.49) or a decrease of at least 7.5% (approximately g ≈ –0.67), based on estimates derived from a hypothetical patient with a baseline femoral neck BMD of 0.80LJ±LJ0.09LJg/cm² [84]. As such, the observed change is unlikely to translate into a meaningful reduction in the increased fracture risk for transgender women compared to both cisgender men and women [64].

Another important limitation in the current body of literature, and consequently in our review is the predominant reliance on DXA as the imaging modality for assessing bone health. While DXA provides clinically useful measures of areal bone mineral density (aBMD), it does not capture bone microarchitecture or strength parameters, which are critical determinants of fracture risk. High-resolution peripheral quantitative computed tomography (HR-pQCT), a more advanced imaging technique, offers superior resolution of trabecular and cortical microstructure and may serve as a better surrogate for fracture risk, particularly in populations with altered bone physiology such as transgender women.

Emerging data from HR-pQCT studies, such as Bretherton et al. [57], suggest that transgender women may exhibit compromised bone microarchitecture compared to cisgender men, potentially highlighting an underappreciated aspect of musculoskeletal health in this population. However, such changes may be influenced by suboptimal oestradiol levels achieved through gender-affirming hormone therapy (GAHT), as indicated by recent findings in humans [63] and mice [85, 86]; underscores the need for more nuanced research into oestradiol dosing, monitoring, and its skeletal effects. Future systematic reviews and primary research should aim to incorporate HR-pQCT data where available and explore the relationship between hormone levels, bone microarchitecture, and fracture risk more comprehensively. Addressing these gaps will be essential for optimising clinical care and bone health outcomes in transgender women.

A key limitation of this meta-analysis is the lack of uniformity across studies regarding the type, duration, and dosing of GAHT, as well as the variable inclusion of gonadectomy status. These factors are known to significantly influence musculoskeletal outcomes, particularly BMD and long-term bone health. Additionally, nutrition, physical activity, differences in age at initiation of GAHT and duration of hormone exposure introduce further heterogeneity, potentially impacting the comparability of findings across studies and limiting the precision of pooled estimates.

### Clinical Implications

The observed beneficial effect of GAHT on FN BMD suggests a protective role in bone health in transgender women, potentially reducing osteoporosis risk. However, the small non-clinically meaningful effect size, the absence of a significant effect on LS and negative trend in TH BMD, the lack of HR-pQCT use, alongside high heterogeneity and low strength of evidence in BMD results, highlights the need to implement improved bone health monitoring in transgender women globally. Notably, there is a lack of empirical data on ageing transgender women above 50 years old, making it unclear whether these trends persist later in life, resulting in practitioners being unsure of how to prescribe elderly transgender women. Additionally, the large effect size increases in FM and BF% and decreases in mCSA and FFM raise concerns about long-term muscle and metabolic health, including heightened risk factors for sarcopenic obesity [87], cardiovascular disease [88] and type 2 diabetes, although increased diabetes incidence has not been found at the population level [89]. Given the absence of detrimental effects on muscle strength, targeted interventions such as resistance training may help mitigate sarcopenic and metabolic disease risk. These findings underscore the need for routine musculoskeletal and metabolic health assessments in transgender women undergoing GAHT in clinical care, particularly as they age.

To enhance clinical management of bone health in transgender women, we recommend routine monitoring that combines DXA to assess areal BMD and body composition, alongside high-resolution peripheral quantitative computed tomography (HR-pQCT) to evaluate volumetric BMD, cortical porosity, and trabecular microarchitecture. This multimodal approach allows a more nuanced understanding of skeletal integrity beyond aBMD alone. Importantly, bone health monitoring should be interpreted in the context of physical activity levels, nutritional status, GAHT prescription type and duration, and gonadectomy status. Future research should prioritise longitudinal studies that integrate these factors to inform targeted bone health interventions and optimise long-term musculoskeletal outcomes in this population.

## Conclusion

This meta-analysis indicates that long-term GAHT in transgender women is associated with increased FN BMD, alongside significant gains in FM and BF% and reductions in FFM and mCSA . Cross-sectional comparisons between transgender women and cisgender men reveal higher fat mass and body fat%, lower fat-free mass and muscle strength, though these differences may reflect pre-existing variations rather than GAHT-induced effects. Given the high heterogeneity in several outcomes and the absence of empirical data on ageing transgender women, further research and clinical monitoring of bone health are needed to clarify the long-term implications of GAHT on bone and musculoskeletal health, particularly as transgender women age.

## Supporting information

Risk Of Bias

Search Strategy

Codebook

Original List of Studies

## Data Availability

All data produced are available online https://osf.io/42wy3/

## Contributions

Conceptualisation, BRH; methodology, BRH; writing-original draft preparation, BRH; Data Abstraction. AB, SMM, BRH; Risk of Bias, AB and SMM; GRADE, BRH; Clinical Applications, JS, LC and JB; Data analysis, BRH; writing--review and editing, ALL.

## Acknowledgements

The authors wish to express their gratitude to Dr Ferus Guppy for their guidance in developing the methodology and to Dr George Kelley for their guidance in the development of the codebook used for data abstraction. The authors would also like to thank Dr Tommy Lundberg for supplying the requested data from Wiik et al (2019).

## Abbreviations

The following abbreviations are used in this manuscript:

BMD: Bone Mineral Density
LM: Lean Mass
FFM: Fat-Free Mass
FM: Fat Mass
mCSA: Muscle Cross-Sectional Area
BMI: Body Mass Index
DPA: Dual-energy Photon Absorptiometry
DXA: Dual-energy X-ray Absorptiometry
ES: effect size
FN: Femoral Neck
TH: Total Hip
G: Hedges standardised mean difference effect size
GRADE: Grading of Recommendations Assessment, Development and Evaluation
IVhet: Inverse heterogeneity
LFK: Luis Furuya-Kanamori
LS: Lumbar Spine
NNS: Number-needed-to-screen
PRISMA: Preferred Reporting Items for Systematic reviews and Meta-Analyses
pQCT: peripheral Quantified Computer Tomography
SD: Standard Deviation

