## Supplemental Figures for "The Effects of Gender Affirming Hormone Treatment on Transgender Women’s Musculoskeletal Health: A Systematic Review and Meta-Analysis"

**a)**

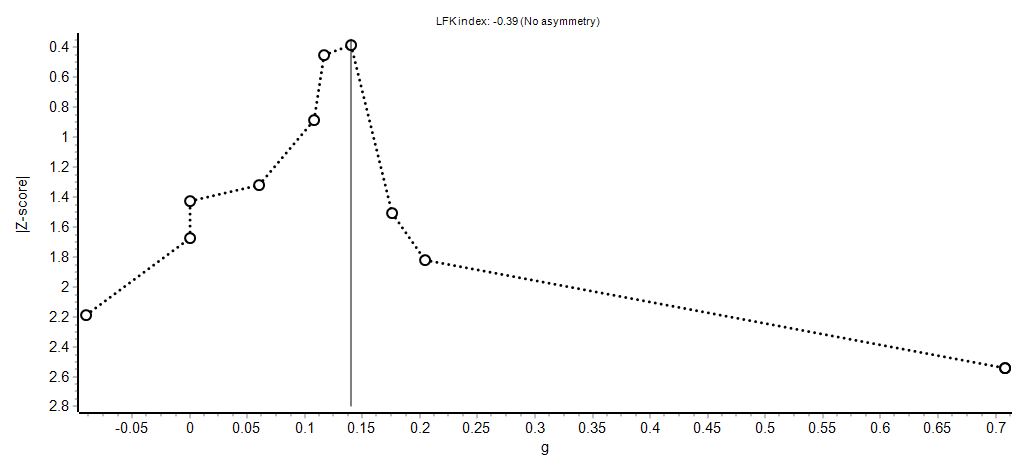

**b)**

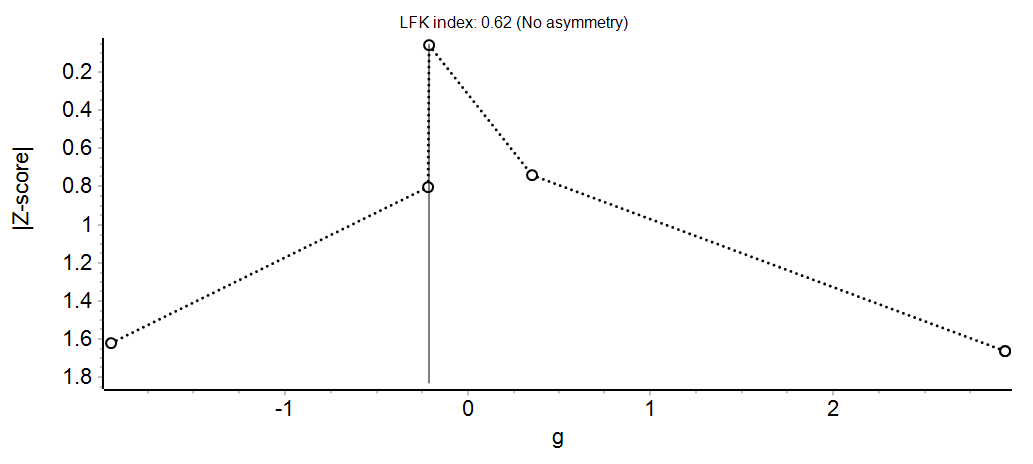

**Supplementary figure 1.** Doi plot for a) longitudinal changes and cross-sectional comparisons in FN BMD with cisgender men. The black circles represent the Z-score and the estimated standardized effect size changes (g) of each effect size (ES) that was assessed for FN BMD the Z-score for each ES. The black solid line represents the pooled g in FN BMD.

**a)**

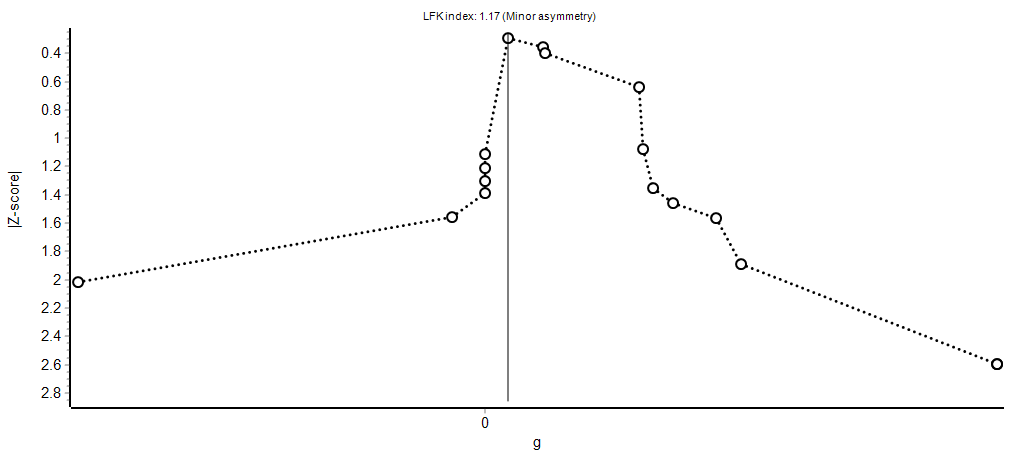

**b)**

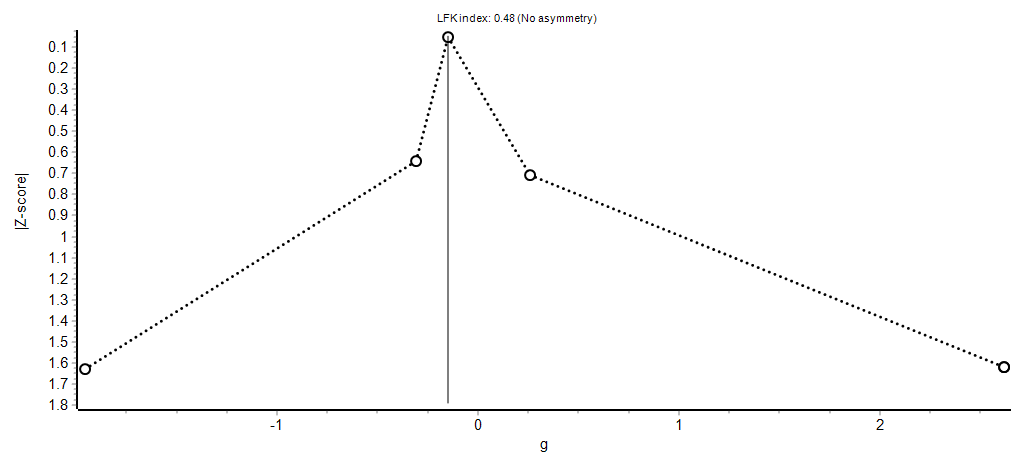

**Supplementary figure 2.** Doi plot for **a)** longitudinal changes In transgender women and **b)** cross-sectional comparisons in LS BMD between cisgender men and transgender women. The black circles represent the Z-score and the estimated standardized effect size changes (g) of each effect size (ES) that was assessed for LS BMD the Z-score for each ES. The black solid line represents the pooled g in LS BMD.

**a)**

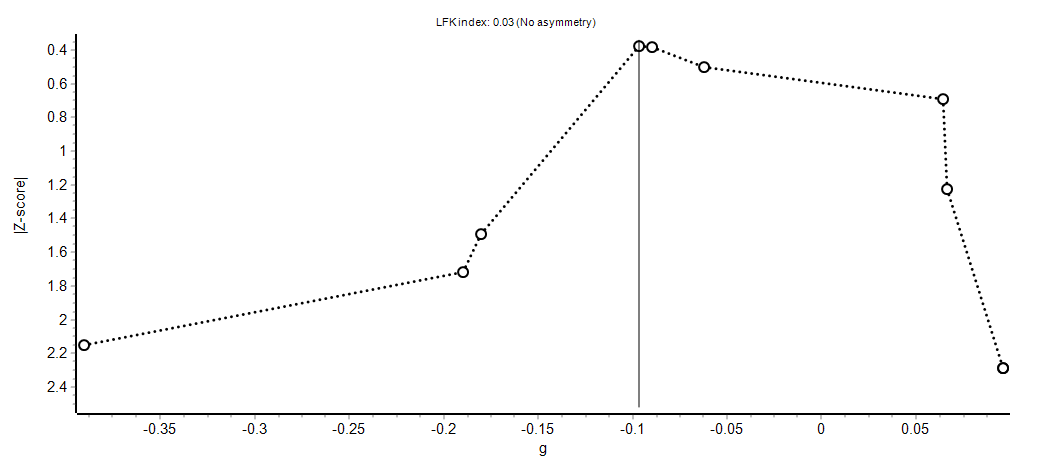

**b)**

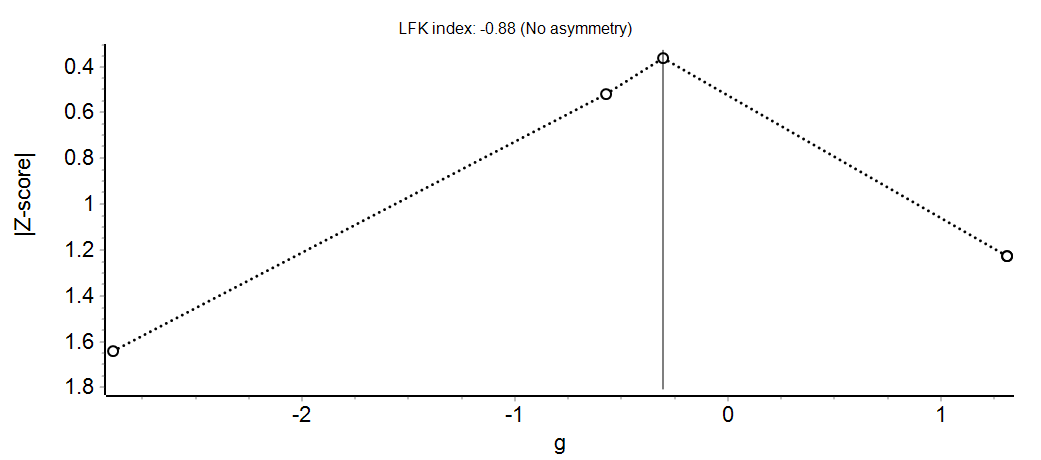

**Supplementary figure 3.** Doi plot for **a)** longitudinal changes In transgender women and **b)** cross-sectional comparisons in TH BMD between cisgender men and transgender women. The black circles represent the Z-score and the estimated standardised effect size changes (g) of each effect size (ES) that was assessed for TH BMD the Z-score for each ES. The black solid line represents the pooled g in TH BMD.

**a)**
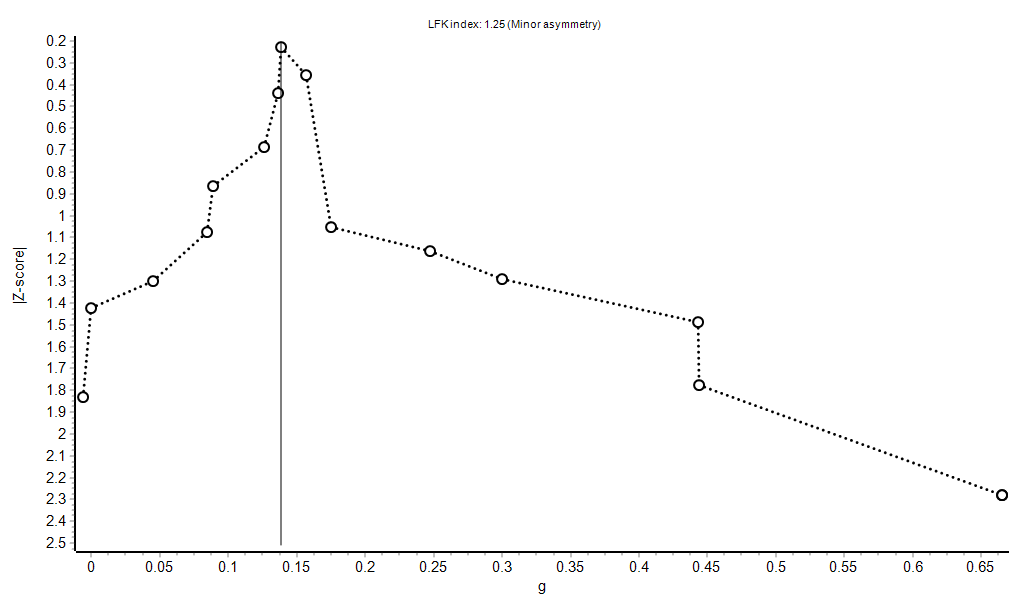

**b)**

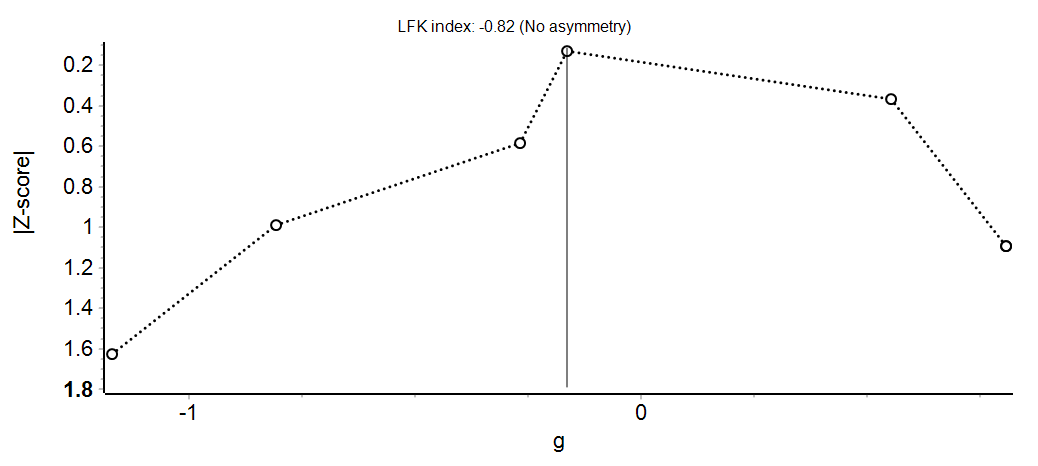

**Supplementary figure 4.** Doi plot for **a)** longitudinal changes In transgender women and **b)** cross-sectional comparisons in BMI between cisgender men and transgender women. The black circles represent the Z-score and the estimated standardised effect size changes (g) of each effect size (ES) that was assessed for BMI the Z-score for each ES. The black solid line represents the pooled g in BMI.

**a)**
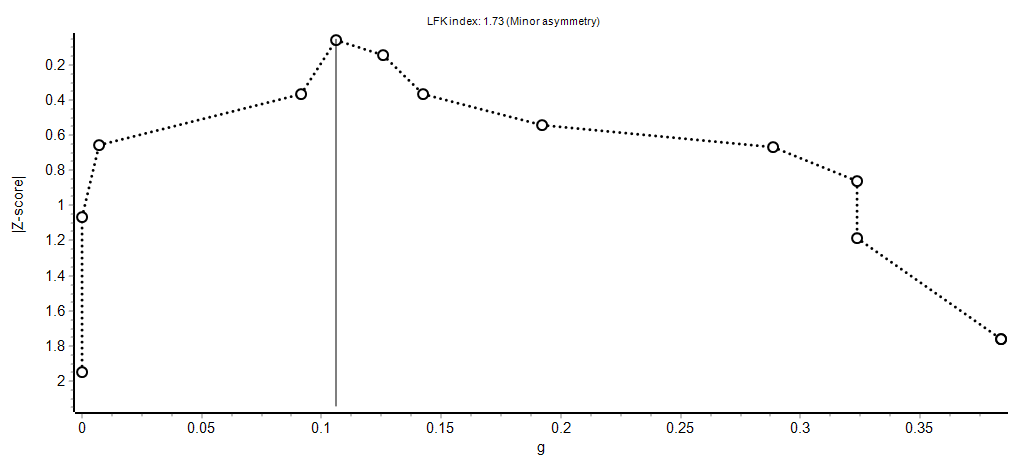

**b)**
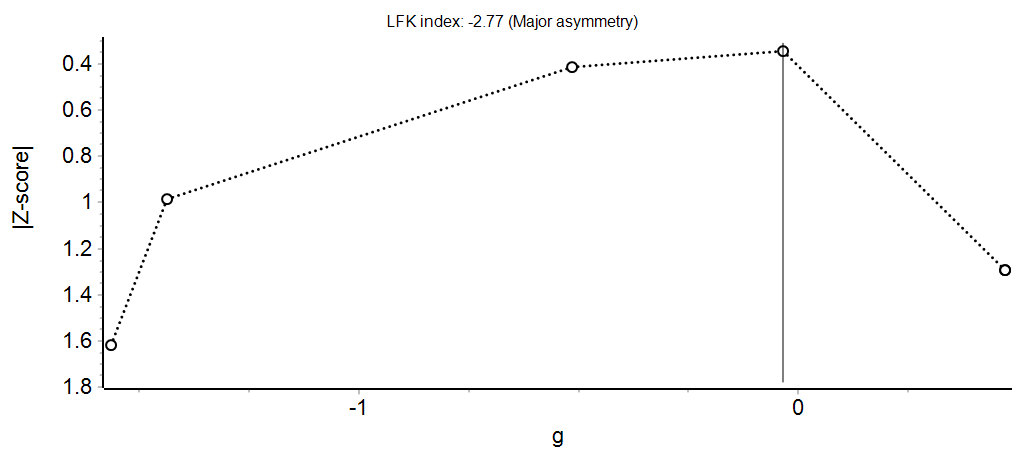

**Supplementary figure 5.** Doi plot for **a)** longitudinal changes In transgender women and **b)** cross-sectional comparisons in Body Mass between cisgender men and transgender women. The black circles represent the Z-score and the estimated standardised effect size changes (g) of each effect size (ES) that was assessed for Body Mass the Z-score for each ES. The black solid line represents the pooled g in Body Mass.

**a)**
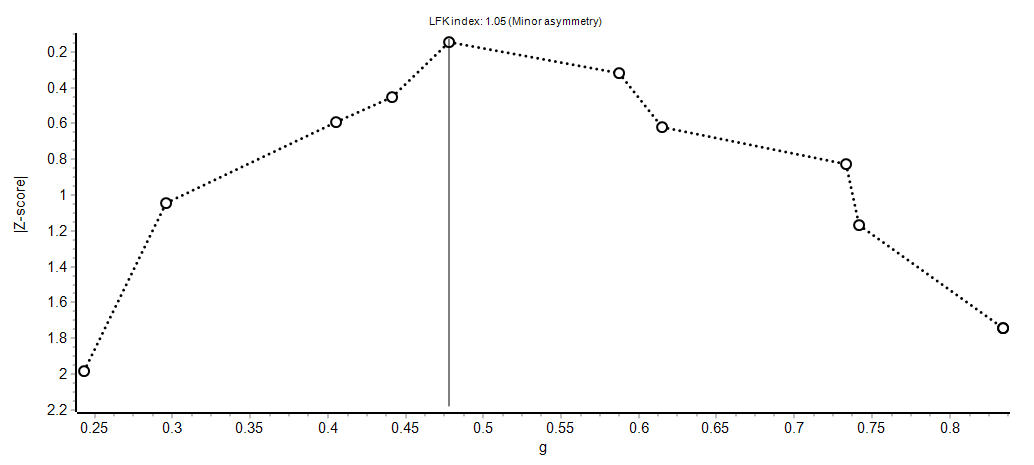

**Supplementary figure 6.** Doi plot for **a)** longitudinal changes In transgender women in Fat Mass between cisgender men and transgender women. The black circles represent the Z-score and the estimated standardised effect size changes (g) of each effect size (ES) that was assessed for Fat Mass the Z-score for each ES. The black solid line represents the pooled g in Fat Mass.

**a)**
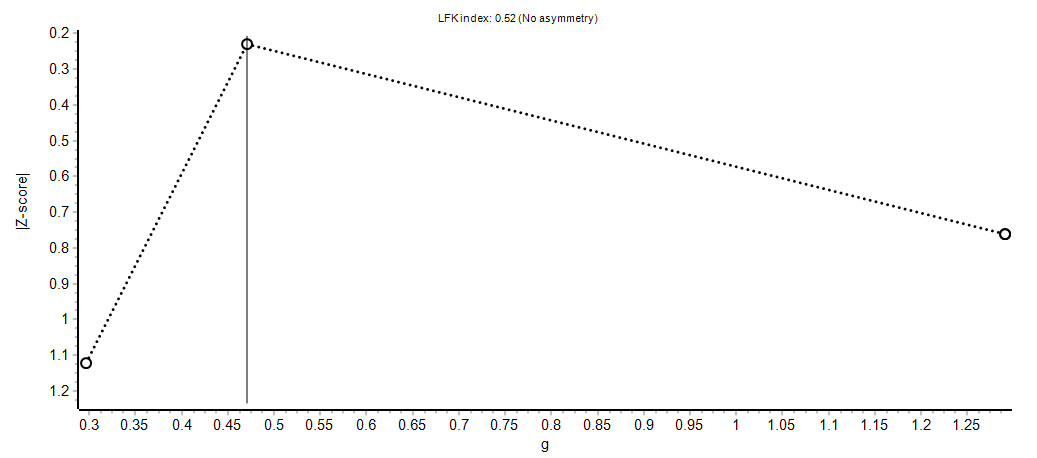

**b)**
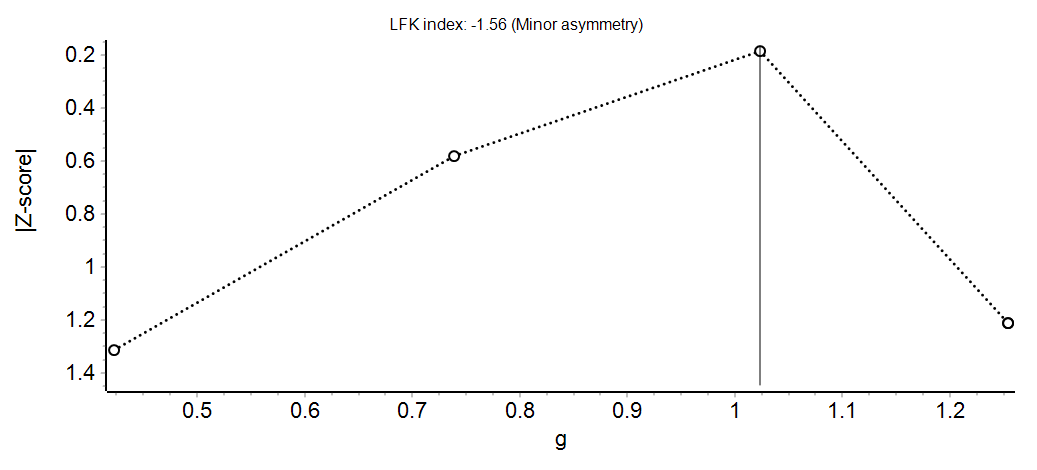

**Supplementary figure 7.** Doi plot for **a)** longitudinal changes In transgender women and **b)** cross-sectional comparisons in Body Fat % between cisgender men and transgender women. The black circles represent the Z-score and the estimated standardised effect size changes (g) of each effect size (ES) that was assessed for Fat Mass % the Z-score for each ES. The black solid line represents the pooled g in Fat Mass %

.

**a)**
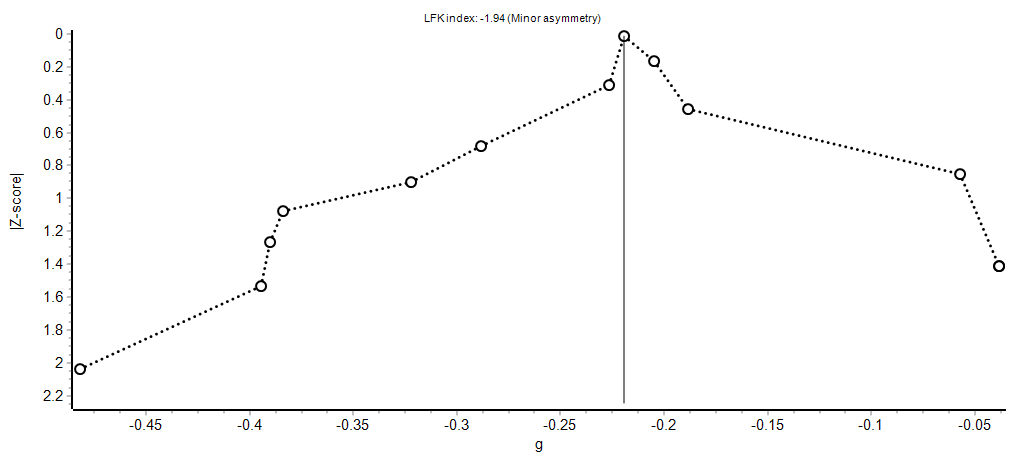

**b)**
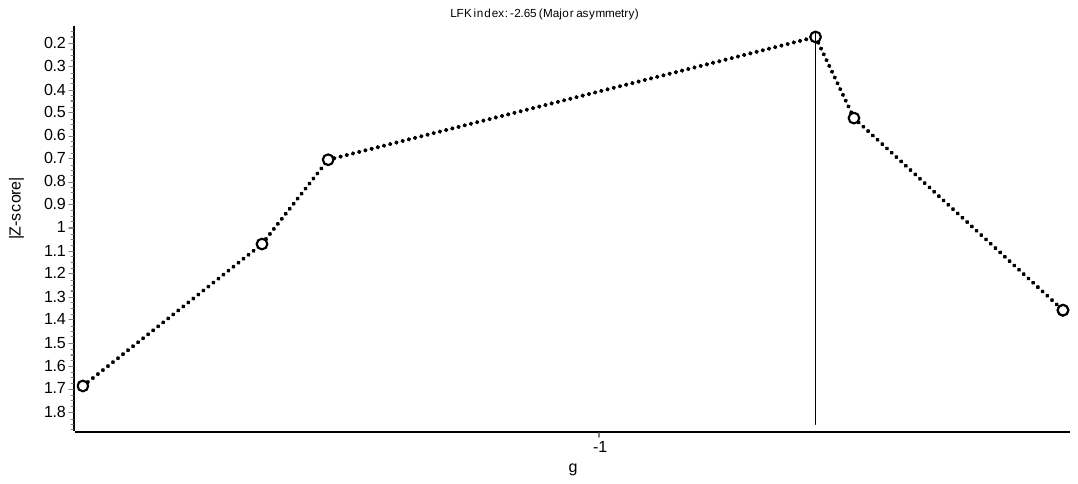

**Supplementary figure 8.** Doi plot for **a)** longitudinal changes In transgender women and **b)** cross-sectional comparisons in *Fat Free Mass* between cisgender men and transgender women. The black circles represent the Z-score and the estimated standardised effect size changes (g) of each effect size (ES) that was assessed for Fat Free Mass the Z-score for each ES. The black solid line represents the pooled g in Fat Free Mass.

**a)**
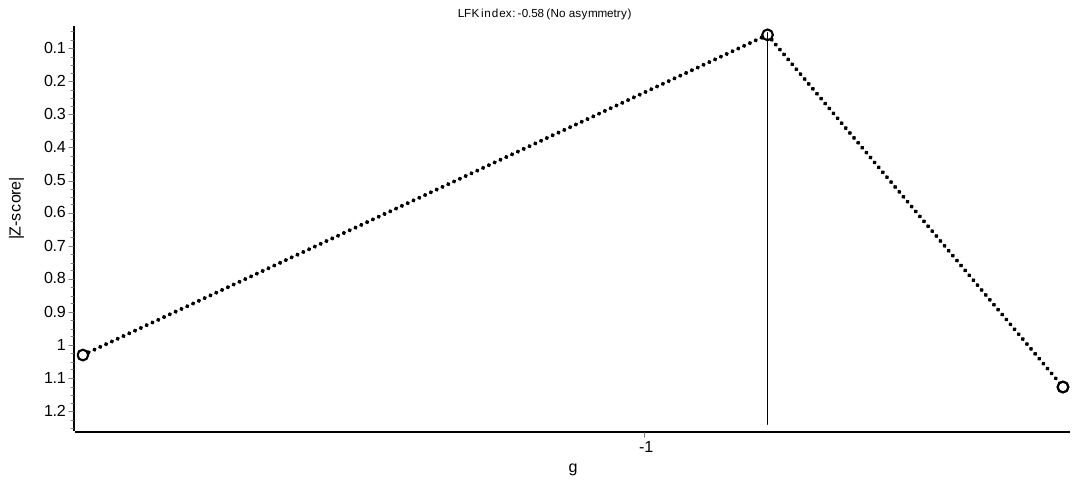

**Supplementary figure 9.** Doi plot for longitudinal changes In transgender women in *Muscle Cross Sectional Area*. The black circles represent the Z-score and the estimated standardised effect size changes (g) of each effect size (ES) that was assessed for Muscle Cross-Sectional Area. The black solid line represents the pooled g in the Muscle Cross Sectional Area.

**a)**
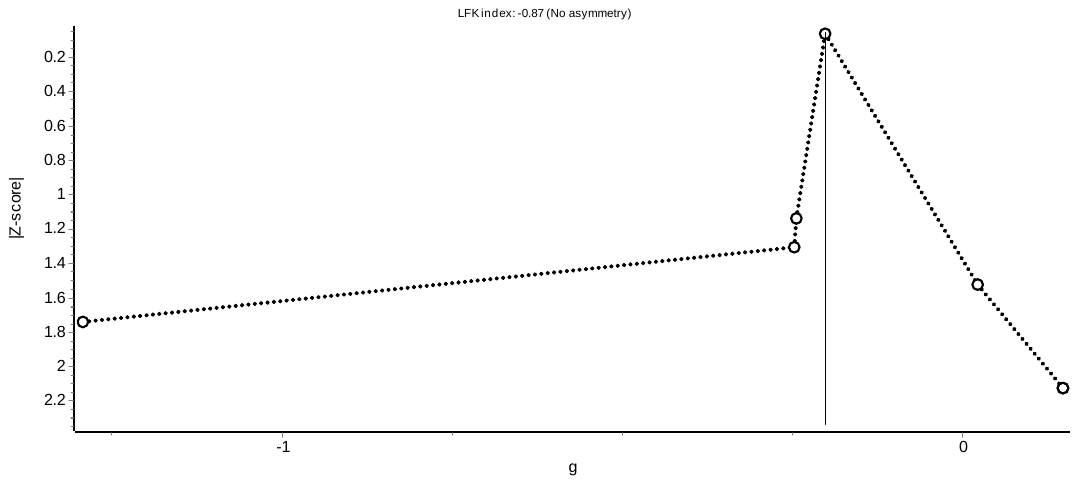

**b)**
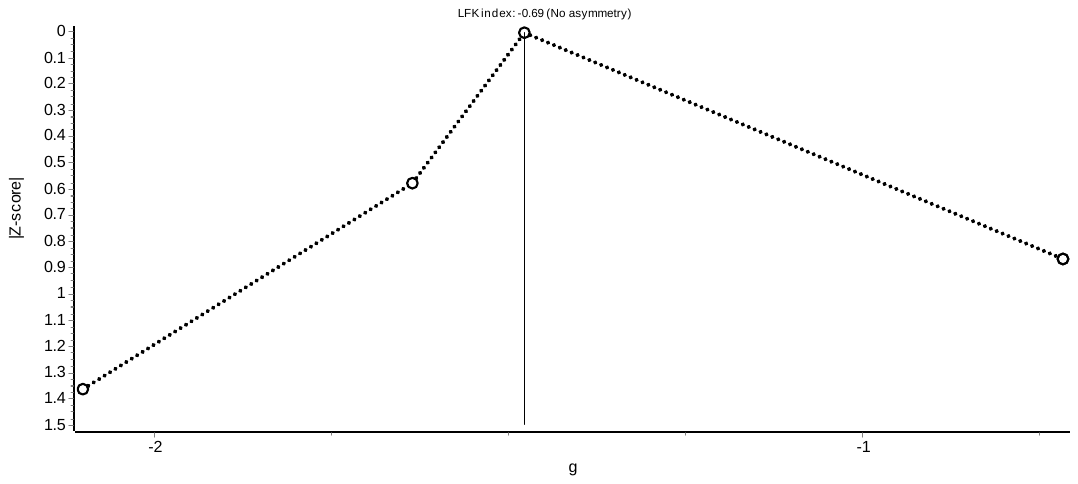

**Supplementary figure 10.** Doi plot for **a)** longitudinal changes In transgender women and **b)** cross-sectional comparisons in *Muscle Strength* between cisgender men and transgender women. The black circles represent the Z-score and the estimated standardised effect size changes (g) of each effect size (ES) that was assessed for Muscle Strength the Z-score for each ES. The black solid line represents the pooled g in Muscle Strength.
