## Supplemental Tables for "The Effects of Gender Affirming Hormone Treatment on Transgender Women’s Musculoskeletal Health: A Systematic Review and Meta-Analysis"

**Supplementary Table 1** Quality of Evidence Assessment

| No. of Studies | Design | RoB | Indirectness of patients, intervention, and comparator | Imprecision | Inconsistency | Publication  Bias | **Quality of Evidence** |
| --- | --- | --- | --- | --- | --- | --- | --- |
| Longitudinal | | | | | | | |
| Femoral Neck | | | | | | | |
| 10 Studies (n=1270) | Observational | Moderate^1^ | Some^2^ | None | None^4^ | None | **Moderate** |
| Lumbar Spine | | | | | | | |
| 16 Studies (n=1594) | Observational | Moderate^1^ | Some^2^ | None | Moderate | Some^5^ | **Low** |
| Total Hip | | | | | | | |
| 11 studies (n=1440) | Observational | Moderate^1^ | Some^2^ | None | None^4^ | None | **Moderate** |
| BMI | | | | | | | |
| 16 Studies (n=645) | Observational | Low^1^ | Some^2^ | None | None^4^ | None | **Moderate** |
| Body Mass | | | | | | | |
| 14 studies (n=279) | Observational | Low^1^ | Some^2^ | Some^3^ | None^4^ | None | **Moderate** |
| Fat Mass | | | | | | | |
| 10 Studies (n=242) | Observational | Low^1^ | Some^2^ | None | None^4^ | None | **High** |
| Body Fat % | | | | | | | |
| 3 Studies (n=47) | Observational | Moderate^1^ | Some^2^ | Some^3^ | Low | None^6^ | **Moderate** |
| Fat-Free Mass | | | | | | | |
| 12 Studies (n=242) | Observational | Low^1^ | Some^2^ | None | None^4^ | None | **Moderate** |
| Thigh Muscle CSA | | | | | | | |
| 3 Studies (n=50) | Observational | Moderate^1^ | Some^2^ | Some^3^ | Moderate | Some^5,6^ | **Moderate** |
| Muscle Strength | | | | | | | |
| 7 Studies (n=313) | Observational | Serious^1^ | Some^2^ | Some^3^ | Moderate | Some^5,6^ | **Low** |
| **Cross-Sectional** | | | | | | | |
| Femoral Neck | | | | | | | |
| 5 Studies (n=233) | Observational | Moderate^1^ | Some^2^ | Some^3^ | Moderate | Some^5,6^ | **Very Low** |
| Lumbar Spine | | | | | | | |
| 5 Studies (n=233) | Observational | Moderate^1^ | Some^2^ | Some^3^ | Moderate | Some^5,6^ | **Very Low** |
| Total Hip | | | | | | | |
| 4 studies (n=180) | Observational | Moderate^1^ | Some^2^ | Some^3^ | Low | Some^5,6^ | **Very Low** |
| BMI | | | | | | | |
| 6 Studies (n=255) | Observational | Critical^1^ | Some^2^ | Some^3^ | Large | Some^5,6^ | **Very Low** |
| Body Mass | | | | | | | |
| 5 studies (n=226) | Observational | Critical^1^ | Some^2^ | Some^3^ | Large | Some^5,6^ | **Very Low** |
| Body Fat % | | | | | | | |
| 4 Studies (n=158) | Observational | Critical^1^ | Some^2^ | Some^3^ | Very Low | Some^5,6^ | **Low** |
| Fat Mass | | | | | | | |
| 2 Studies (n=73) | Observational | Critical^1^ | Some^2^ | Some^3^ | Very Low | - | **Low** |
| Fat-Free Mass | | | | | | | |
| 6 Studies (n=264) | Observational | Critical^1^ | Some^2^ | Some^3^ | Low | None^6^ | **Low** |
| Muscle Strength | | | | | | | |
| 4 Studies (n=116) | Observational | Critical^1^ | Some^2^ | Some^3^ | Moderate | None^6^ | **Low** |
| ^1^ Assessed via the RoBinS II tool.  ^2^ The authors conclude that the differing regimens of GAHT will affect comparators due to differing standards of care across studies.  ^3^ The authors consider there to be some imprecision due to the large confidence intervals in effect size  ^4^ The authors consider the inconsistency of results to be none due to the *I^2^* value being 0 for the outcome.  outcome.  ^5^ Funnell Plot Analysis Showed the presence of one or more study outliers.  ^6^The number of studies for this outcome was below the recommended 10 studies to test for publication bias through the funnel plot method (Sterne et al., 2011) | | | | | | | |

**Supplementary Table 2** Influence Analysis for FN BMD

| **Study** | **Group Excluded** | **Mean (95% CI)** | **Z (p)** | **Q (p)** | **I^2^ (%)** | **LFK Index** |
| --- | --- | --- | --- | --- | --- | --- |
| All | None | 0.13 (0.05, 0.21) | **3.19 (0.00)*** | 4.32 (0.89) | 0 | -0.73 (None) |
| Wiepjes et al (2018) | Transgender Women | 0.11(-0.01, 0.23) | 1.82 (0.14) | 4.19 (0.84) | 0 | 0.15 (None) |

Unless noted otherwise, all outcomes are reported as standardized effect size (g); ES, effect size; #, number; participants (#), number of exercise and control participants nested within ES's and studies; Z(p), z-score and alpha value; Q(p), Cochran's Q statistic and alpha value; I^2^ (%), I-squared.

⁎ Statistically significant (p<0.05).

**Supplementary Table 3** Influence Analysis for Muscle CSA

| **Study** | **Group Excluded** | **Mean (95% CI)** | **Z (p)** | **Q (p)** | **I^2^ (%)** | **LFK Index** |
| --- | --- | --- | --- | --- | --- | --- |
| All | None | -0.48 (-0.90,-0.06) | -2.24 (0.03)* | 19.13 (0.00) | 69 | -3.12 (Major) |
| Elbers et al | Transgender Women | **-0.44 (-0.92, 0.04)** | **-1.81 (0.07)** | 17.97 (0.22) | 72 | 3.00 (Major) |

Unless noted otherwise, all outcomes are reported as standardized effect size (g); ES, effect size; #, number; participants (#), number of exercise and control participants nested within ES's and studies; Z(p), z-score and alpha value; Q(p), Cochran's Q statistic and alpha value; I^2^ (%), I-squared.

⁎ Statistically significant (p<0.05).

**Supplementary Table 4** Influence Analysis for Muscle Strength

| **Study** | **Group Excluded** | **Mean (95% CI)** | **Z (p)** | **Q (p)** | **I^2^ (%)** | **LFK Index** |
| --- | --- | --- | --- | --- | --- | --- |
| All | None | -0.27 (-0.91, 0.39) | -0.82 (0.41)* | 55.52 (0.00) | 75 | -0.87 (None) |
| Chiccarelli et al | Transgender Women | **-0.18 (-0.34, -0.02)** | **-2.17 (0.03)** | 1.10 (0.89) | 0 | 2.47 (Major) |

Unless noted otherwise, all outcomes are reported as standardized effect size (g); ES, effect size; #, number; participants (#), number of exercise and control participants nested within ES's and studies; Z(p), z-score and alpha value; Q(p), Cochran's Q statistic and alpha value; I^2^ (%), I-squared.

⁎ Statistically significant (p<0.05).

**Supplementary Table 5** Influence Analysis for Muscle Strength

| **Study** | **Group Excluded** | **Mean (95% CI)** | **Z (p)** | **Q (p)** | **I^2^ (%)** | **LFK Index** |
| --- | --- | --- | --- | --- | --- | --- |
| All | None | -0.33 (-0.81, 0.15) | -1.36 (0.17)* | 55.52 (0.00) | 73 | 0.03 (none) |
| Scharff and Wiepjes | Transgender Women | **-0.47 (-0.88, -0.06)** | **-2.27(0.02)** | 36.48 (0.00) | 73 | 1.70 (Minor) |

Unless noted otherwise, all outcomes are reported as standardized effect size (g); ES, effect size; #, number; participants (#), number of exercise and control participants nested within ES's and studies; Z(p), z-score and alpha value; Q(p), Cochran's Q statistic and alpha value; I^2^ (%), I-squared.

⁎ Statistically significant (p<0.05).
